# A Flexible and Responsive Remote Study Design to Assess Gene Expression Changes During Wildfire Smoke Exposure with homeRNA, an At-home Blood Sampling Kit

**DOI:** 10.1101/2025.10.11.25337783

**Authors:** A. J. Haack, L. G. Brown, Y. Zeng, T. Khan, I. H. Robertson, D. S. Kennedy, K. N. Adams, J. W. MacDonald, T. K. Bammler, F. Stefanovic, K. Moloney, J. E. Stolarczuk, M. G. Takezawa, M. Y. Alizai, G. W. Hassan, F. Y. Lim, D. Chaussabel, E. G. Walker, N. A. Errett, E. Berthier, A. B. Theberge

## Abstract

Transcriptomic responses to wildfire smoke are difficult to study given the unpredictability of wildfires and the challenges of collecting blood during active disasters. To overcome these challenges, we developed a flexible study design leveraging homeRNA, our at-home blood collection and RNA stabilization kit. Between June 2021 and April 2022, 58 participants across 10 U.S. states collected 635 blood samples before, during, and after wildfire events. This responsive approach captured three exposure groups: high exposure in Okanogan County, Washington, medium exposure from transported smoke, and low exposure. During the 10-month study, 93% of participants (n=54/58) returned at least 6 samples. In a preliminary exploratory analysis, we analyzed 770 genes with a Nanostring panel from nine participants (6 high, 3 low-medium exposure) using the BloodGen3 framework. In the high exposure participants, we observed trends toward overexpression of inflammation (inflammation aggregates A33 and A35, and modules M13.1 and M13.12), with concurrent underexpression of adaptive immune responses (lymphocytic aggregates A1 and A6, B cell module M13.18, T cell modules M16.24 and M15.38). This study establishes that homeRNA enables flexible, responsive sampling during disasters, overcoming traditional logistical barriers to capture time-sensitive biological data across dispersed populations.

## INTRODUCTION

The incidence and severity of wildfires have become a global concern, with many regions experiencing increases in fire season lengths and fire intensity.^1–3^ A key pollutant present in wildfire smoke is PM_2.5_, defined as particulate matter 2.5 μm in diameter or smaller, which comprises about 90% of wildfire smoke.^4–6^ Between 2007 to 2018, wildfires have accounted for about 25% of PM_2.5_ concentrations across the United States (U.S.) and up to 50% in the Western U.S.^7,8^ Even in states where there are no wildfire burns in a given year, smoke from wildfires in other states or Canada can exacerbate PM_2.5_ exposure due to transported smoke.^9^ Wildfires are also increasingly prevalent globally, with major impacts documented in Australia, India, Indonesia, Brazil, Mediterranean Europe, Canada, and across African savannas.^10,11^

Wildfires not only have a severe impact on the environmental landscape but also are a contributor to adverse human health outcomes.^12–19^ With the growing body of literature on the effects of wildfire smoke exposure on overall health, there has also been a push to understand the impacts of PM_2.5_ exposure from a biomolecular mechanistic perspective, particularly on the respiratory tract and systemic inflammation.^20–23^ Understanding the biological mechanisms involved in wildfire smoke exposure could enable biomarker discovery, targeted therapy development, and development of risk assessment tools for smoke exposure prevention and response. Moreover, understanding the transient immune response to wildfire smoke is critical as it can affect medical interventions ranging from vaccine efficacy^24^ to reproductive health outcomes including sperm quality^25,26^, potentially influencing both the timing and distribution of healthcare services during smoke events.

There have been multiple studies that have investigated acute inflammatory activation in wildland firefighters in response to occupational smoke exposure.^20,23,27–31^ Most studies that capture wildfire smoke effects in the general population (i.e., smoke events unrelated to occupational exposure or controlled burns) are retrospective, where they take advantage of a separate study that happens to occur while a wildfire happens. Recently, Aguilera et al. designed a study to capture immune response to wildfire smoke exposure, where participants in an urban center came in for phlebotomy draws before, during, and after wildfire events. This study also found alterations in inflammatory protein prevalence in human blood in response to wildfire smoke.^32^ Johnson et al. recently examined immune responses in smoke-exposed individuals, demonstrating increased activation markers on memory T cells via mass cytometry and identifying 133 differentially methylated gene loci associated with smoke exposure^33^ adding to other studies that have investigated the effects of wildfire smoke on DNA methylation^34,35^, but there have been few studies that have investigated the transcriptomic immune response to wildfire smoke exposure in humans.

Understanding why such transcriptomic studies are rare requires examining the unique methodological challenges of wildfire smoke research. Typically, in person clinic-based blood draws are used to study longitudinal immune responses in blood transcriptomes. However, a similar study design for investigating the effects of wildfire smoke exposure is logistically challenging due to the unpredictable nature of wildfires and the low accessibility to healthcare and research centers in rural areas, where many fires occur. Running a clinic-based blood draw study on the general population experiencing wildfire smoke exposure would require either (1) relying on historical data to choose a location that has the infrastructure to support frequent clinic-based blood draws and a high prevalence of wildfire smoke^32^, (2) setting up a multi-site clinic-based study to increase the chances that at least one site has a wildfire smoke event, or (3) relying on retrospective studies or capturing smoke exposure incidentally on a study investigating a different question where longitudinal blood samples are collected.^24,35,36^ These factors make designing studies where blood is collected at multiple time points in response to wildfire smoke exposure logistically challenging and limit the number of participants included from rural areas.

To address this challenge, we employed a remote and flexible study design using homeRNA: a self-sampling kit comprising a commercially available blood collection device, Tasso-SST, and a custom engineered RNA stabilization tube we previously developed.^37^ The homeRNA kit allows study participants to self-collect and stabilize blood by themselves in their own homes for shipment back to a centralized lab for downstream analysis. We have already demonstrated the ability to use homeRNA in high temperature settings^38,39^ and to assess gene expression changes during acute respiratory infections from SARS-CoV-2.^40,41^ Here, our primary objectives were to (1) evaluate the feasibility of a study design that allowed flexible responsive sampling during disasters, (2) demonstrate the usability of homeRNA across a 10-month longitudinal study, and (3) assess the immediate and longitudinal effects of wildfire smoke exposure on gene expression in an initial cohort.

By facilitating self-sampling at home, homeRNA allows for a study design that includes a wider reach of participants living in remote locations who are traditionally inaccessible by mobile phlebotomists or clinic-based blood draws. Study kits can be mailed to participants within a day of a disaster warning or occurrence (such as a wildfire event), thus reducing the need to predict where the disaster would occur ahead of the study as would be required for a clinic-based blood draw study design. In contrast, in-clinic appointments need to be scheduled in advance and may not occur until days or up to a month after exposure, thereby missing the timescale of response that may be more immediate. Furthermore, multiple kits can be sent in one shipment that can be used by participants over several time points upon receiving, allowing for repeated sampling across different groups in response to an event as well as within a single individual throughout a repeated exposure. Here, we implemented homeRNA into a wildfire smoke exposure study to collect blood from 58 participants (635 total samples) across the Western and South Central U.S. before, during, and after wildfire smoke exposure from June 2021 to April 2022. We analyzed a subset of these samples with a targeted Autoimmune response gene panel followed by a gene-set enrichment visualization and interpretation repertoire known as BloodGen3. We analyzed high wildfire exposure response in six participants located in Okanogan County, Washington, which experienced two major wildfires during this time frame that resulted in Air Quality Index (AQI) categories ranging between very unhealthy (PM_2.5_ 125.5 - 225.4 µg/m^3^) and hazardous (PM_2.5_ ≥225.5 µg/m^3^) for two weeks in July 2021. We compared the immune response in participants who were being exposed during these dates (n=6) with the immune status of participants located elsewhere (California (n=2) and Western Washington (n=1)) during the same timeframe. The participants not located in Okanogan County (n=3) experienced low and moderate wildfire exposure throughout the study, with AQI categories ranging between good (PM_2.5_ ≤ 9.0 µg/m^3^) to unhealthy (PM_2.5_ 55.5 - 125.4 µg/m^3^) throughout the season.

## MATERIALS & METHODS

### homeRNA kit components

homeRNA is a self-sampling kit for the collection and stabilization of whole blood RNA.^37^ The kit uses a commercially available Tasso-SST™ blood collection device for the self-collection of peripheral blood and a specially engineered stabilizer tube containing RNA*later*™ for immediate stabilization of cellular RNA. The Tasso-SST™ blood collection device was purchased from Tasso, Inc (Seattle, WA). The stabilizer tube was injection molded out of polycarbonate (PC: Makrolon 2407) by Protolabs, Inc (Maple Plain, MN) and was designed to hold an RNA stabilization solution and connect to the Tasso-SST™ blood collection tube. The stabilizer tube was filled with 1.4 mL RNA*later*™ (Thermo Fisher) stabilization solution and capped. The stabilizer tube insert was injection molded out of polycarbonate (PC: Makrolon 2407) by Protolabs, Inc (Maple Plain, MN) and was designed to hold the stabilizer tube containing blood in a 50 mL conical tube during shipping. All components were cleaned via sonication in 70% ethanol (v/v) for 30 min and air dried prior to assembly. Finally, all kit components were placed in a rigid custom-designed mailer box (The BoxMaker, Inc.). Detailed design files and the workflow of assembling and using the homeRNA kit can be found in an initial pilot study in which we characterized the feasibility of using homeRNA for remote blood collection and RNA stabilization and included in the Supporting Information of this manuscript fro ease of reference.^37^

### homeRNA kit use workflow

The workflow of the homeRNA kit is described in detail in Haack, Lim, et al.^37^ Briefly, to use the homeRNA kit, participants first collected capillary blood from their upper arm using the Tasso-SST™ device. The authors note that the serum separator tube (SST) gel (included in the Tasso-SST collection tube) is not necessary for RNA stabilization and analysis. At the time of the study, the Tasso-SST was the only available Tasso device for purchase. Next, participants were asked to estimate the volume of the blood collected based on a reference photo of the Tasso- SST™ tube in the sample collection survey (Figure S1C). The participants then stabilized the blood in RNA*later*™ by interfacing the RNA stabilizer tube with the Tasso-SST™ blood tube and shaking vigorously to mix the blood with the stabilizing solution. Finally, after mixing, the stabilized blood sample was placed inside a 50 mL conical tube containing an insert to hold the sample tube in place, which was then packaged within a custom homeRNA cardboard box and UPS LabPak and mailed back to the lab for downstream analysis.

### Participant recruitment and enrollment

The study was approved by the University of Washington (UW) Institutional Review Board (IRB) under protocol STUDY00012463. All study procedures were performed after informed consent was obtained. All samples were collected remotely by study participants and online surveys were administered through Research Electronic Data Capture (REDCap). Participants were recruited via social media, community outreach (e.g., through Clean Air Methow, a community air quality program^57,58^), and word of mouth between June and September 2021. After completing a screening survey online, individuals were invited to complete the informed consent and enrollment by the study team. To be eligible for the study, participants had to be: (1) 18 years old or older; (2) living in an area prone to wildfire including the states of Alaska, Arizona, California, Colorado, Florida, Idaho, Montana, Nevada, New Mexico, Oklahoma, Oregon, Texas, Utah, Washington, and Wyoming; (3) not pregnant (upon enrollment); (4) not residing in a correctional facility; and (5) not a friend or family member of researchers conducting study. Participants were enrolled with preference given to: (1) those located in Okanogan County, Washington (WA) (a historically wildfire-prone area) and (2) those with diverse geographic locations outside of Okanogan County.

### Sample collection timeline

Throughout the study, participants were sent a package containing three homeRNA kits, and given a schedule for when to use each kit. Typically, each of the three samples was taken 3 to 4 days apart, and returned immediately via overnight shipping, if available. Depending on the smoke exposure conditions, participants were sometimes sent an additional set of three kits if there was continued wildfire smoke exposure in their area.

### Before wildfire season

Immediately upon enrollment, participants were sent three samples to serve as baseline time-point samples before the start of wildfire season (June 2021). Samples were collected 2-4 days apart from each other.

### During wildfire season

The location of wildfires, smoke plumes, and PM_2.5_ values were monitored throughout the wildfire season (July-September 2021). If smoke was present in a participant’s area, participants were immediately sent a box with three kits and asked to sample every 2-4 days. During the wildfire smoke events in Okanogan County, WA, participants outside of this region who were not experiencing smoke exposure were also asked to sample at similar times to match participants located in Okanogan County.

### After wildfire season

All participants were sent an additional set of three homeRNA kits between October - November 2021 (3 months after the wildfire events in Okanogan County), then again in late February - early April 2022 (approximately 6 months after the wildfire events in Okanogan County). For each timepoint, samples were collected every 2-4 days.

### PM_2.5_ data collection

The average daily concentration of PM_2.5_ that each participant was exposed to during the study period was taken from each participant’s nearest outdoor EPA PM_2.5_ monitor. A subset of participants were also provided an indoor PurpleAir monitor to collect indoor PM_2.5_ data

*Outdoor PM_2.5_*

Daily average PM_2.5_ data collected by the closest EPA PM_2.5_ monitor to each participant’s home address was downloaded from the EPA website.^59^ The closest monitor was determined by using the equirectangular approximation to calculate the shortest distance between the coordinates of each participant’s home address and the reported coordinates of EPA PM_2.5_ monitors. If no average PM_2.5_ data were available on a certain day with a participant’s calculated nearest monitor, then PM_2.5_ data were taken from the calculated second nearest EPA PM_2.5_ monitor. If the second nearest EPA monitor did not have available data, then data were taken from the calculated third nearest PM_2.5_ monitor. For a few cases, there were no data from the nearest three monitors, and these particular days were excluded from the analysis.

#### Indoor PM_2.5_

A subset of participants (n=15) were provided with indoor PurpleAir monitors, and the study team was provided with the sensor ID. Indoor PM_2.5_ data collected by participant’s PurpleAir sensors were downloaded from the PurpleAir website^60^, and the daily average PM_2.5_ from the PurpleAir sensor was calculated for each day of the study period.

### Exposure level categorization

Participant exposure levels were categorized as high, medium, and low based on the standard deviation of the mean of their total outdoor PM_2.5_ values throughout the study period (June 2021 - April 2022). Those with a standard deviation >20 µg/m^3^were considered high exposure participants; those with a standard deviation between 10 and 19 µg/m^3^ were considered medium exposure participants; those with a standard deviation <10 µg/m^3^ were considered low exposure participants. Given that ambient air quality was generally good throughout most of the study period, with PM_2.5_ spikes concentrated during the 3 month wildfire season, we used standard deviation rather than mean PM_2.5_ as our primary metric for classifying participants into exposure groups. This approach specifically identifies individuals who experienced high-magnitude episodic exposures, which would be obscured by time-averaged means in a population with uniformly low baseline exposures.

### Survey data collection

Participants were surveyed throughout the study period through REDCap. Specific surveys are described below and all surveys with corresponding questions are available in the Supporting Information:

### Enrollment Survey

This survey was given immediately after informed consent was acquired. It collects information about participant demographics, home addresses (for sending samples and tracking PM_2.5_ data), as well as underlying health conditions, recent vaccinations, and medications that may affect immune activation.

### Baseline Survey

This survey was given with the first blood sample collected, whether it was before the first wildfire smoke exposure or not. The baseline survey collects information on exposures a participant may have experienced in the past year, the participant’s occupation (specifically if their profession involves working outside), and the participant’s typical daily activities. Questions on daily activities and stress were adapted from standardized surveys on stress assessment available on PhenX.^61^ The baseline survey also includes all the questions in the Sample Collection Survey (below), and if the participant was experiencing smoke exposure for the first sample, they were asked additional questions that are specific to smoke exposure listed in the Smoke Exposure Survey (below).

### Sample Collection Surveys

This survey was given with every blood sample collected. It includes questions about the date and time of the sample collection, sample collection metrics (e.g., blood volume estimation, time used for the collection), and homeRNA usability (e.g., ease of use with the kit, pain level using the Tasso device). The survey also included questions about any symptoms related to smoke exposure (the list of symptoms was adopted from a CDC website on health effects of wildfire smoke^62^), they were asked to rate their stress levels compared to normal, they were asked to estimate hours of sleep, and they were asked about whether or not they were able to engage in their normal daily activities. If a participant received an indoor PurpleAir monitor, they were asked to report the PM_2.5_ value on the air monitor at the time of collection.

### Smoke Exposure Surveys

This survey was given if a study participant was using the homeRNA kit during wildfire smoke exposure. It contains all the questions in the Sample Collection Survey (above) with additional questions specifically about behaviors in response to wildfire smoke exposure. These questions included how much time the participants spent outdoors, in a car or bus, and the amount of time spent indoors. It also asked if they utilized any mitigation strategies, such as using an air filter, wearing masks, avoiding outside activities, or keeping windows closed.

### Closing Survey

This survey was given at the end of the study period. It included demographic and behavior questions, including if a participant had a major change in behavior, health status, or location (e.g., on vacation) during the study.

### Follow-up Survey

This survey was given in November 2023, which was 18 months after the study was concluded in April 2022. Of the 40 participants invited to complete the survey, 30 participants provided responses and completed the follow-up survey. This survey asked for the participant’s motivation to enroll in the study, comparisons between remote blood sampling and in-person blood draws, feedback on using the homeRNA kit, and willingness to participate again.

### Sample processing

After the homeRNA kit was mailed back to the lab, it was stored immediately in a -20°C freezer on campus. Within a week, the 50 mL conical tube containing the blood tube connected to the stabilizer tube (which contained the homeRNA-stabilized blood sample) was removed from the kit and transferred to -80°C until ready for further processing. Total cellular RNA was isolated from samples using the Ribopure™ - Blood RNA Isolation Kit (Thermo Fisher) according to the manufacturer’s protocol and eluted in two 50 μL aliquots. Isolated RNA was stored at -80°C until ready for further analysis.

### Assessment of total cellular RNA integrity and yield

The RNA Integrity Number (RIN) scores of the first 50 μL elution were obtained on a Bioanalyzer 2100 (Agilent) using the Agilent RNA 6000 Pico Kit (Agilent 5067-1513). All RNA samples were diluted 1:20 in nuclease-free water before RIN measurement to ensure the RNA concentration of the loaded sample was in the qualitative range for the Agilent RNA 6000 Pico Kit. The RNA concentrations of the first 50 μL elution were measured using the Qubit Flex Fluorometer (Thermo Fisher) using the Qubit RNA High Sensitivity Assay Kit (Invitrogen Q32855).

### Participant and sample selection for Nanostring analysis

For an initial exploratory analysis using a Nanostring panel, nine participants were selected from the total 58 participants who completed the study. As Okanogan County experienced the most profound smoke exposure during the study period, six of the nine selected participants were located in this region. We also selected three additional participants; one was located in central Washington who experienced a moderate amount of smoke exposure due to their proximity to Okanogan County, and the other two were located in California, who experienced little to no smoke exposure in the 2021 wildfire season.

Participant selection preference was given to participants if they had (1) at least five samples collected with associated surveys and if (2) at least one of the five samples collected was collected during wildfire smoke exposure (if located in Okanogan County) or with a similar time point (if not located in Okanogan County). In total, we submitted 107 samples from nine participants for gene expression analysis using the nanoString nCounter Autoimmune Profiling Panel. Additional RNA processing was performed on a subset of these samples; these RNA samples were treated with DNase I (RNase-free) (New England Biolabs, M0303S) to remove contaminating genomic DNA and/or Monarch® RNA Cleanup Kit (New England Biolabs, T2030L) to purify and concentrate the RNA according to the manufacturer’s protocols. The final cutoff RIN value for samples after processing was >6. Most samples used an input of 100 ng, but a handful of samples had inputs less than 100 ng. Input volumes ranged from 8-10 μL.

### Gene Expression Analysis

#### nCounter data quality control and normalization

Raw nCounter RCC files were processed using custom Python scripts. Quality control metrics were evaluated for all samples prior to normalization following established nCounter guidelines^63,64^. Imaging quality required a minimum of 75% fields of view (FOV) successfully registered, while binding density was assessed with acceptable ranges of 0.05-2.25 spots/μm². Positive control linearity was evaluated through correlation analysis with expected synthesis RNA concentrations (0.125-128 fM), requiring R² > 0.95 to ensure proper hybridization efficiency. Negative control probes were assessed for background levels, with mean plus two standard deviations used as detection thresholds. Housekeeping gene variability was evaluated using coefficient of variation, with sample showing CV > 50% flagged for potential exclusion due to high technical variability.

Gene expression normalization was performed using a sequential pipeline combining standard nCounter procedure with additional variance stabilization methods. The process began with positive control normalization utilizing the geometric mean of synthetic positive control probes to adjust for technical variation in hybridization and detection across samples. Codeset content normalization followed, employing eight reference genes (*TUBB, MRPS7, TBP, SDHA, GUSB, HRPT1, NMT1, PGK1*) selected based on low variance and moderate expression levels across the dataset. Background subtraction was performed using negative control statistics, with gene showing expression below detection threshold flagged accordingly. Following these standard steps, data underwent counts per million (CPM) transformation to normalize for library size differences, then trimmed means of m-values (TMM) normalization with 30% trimming to address compositional bias between samples. Final normalized values were log2-transformed for downstream analyses. All data processing was performed using custom Python scripts with numpy (v1.22.3) and pandas (v.1.4.2) libraries.

#### Differential expression analysis

To control for intra-individual baseline variation, participant-specific median expression values for each gene were calculated from pre-exposure baseline samples (timeline category "before") and subsequently subtracted from exposure timepoint measurements corresponding to that participant. This yielded baseline-corrected gene expression values that reflect changes relative to each participant’s pre-exposure state. At the individual gene level, we identified 253 genes with differential expression (FDR < 0.05) between exposed and non-exposed groups. To determine biological significance, we also performed a minimum-effect test where the null hypothesis was |log₂FC| ≤ 0.5.^50^ No genes achieved FDR < 0.05 using this more stringent approach, indicating that while statistical differences exist, no genes showed fold changes greater than 1.4-fold with controlled false discovery rate.

### Gene set enrichment of wildfire smoke exposure samples using BloodGen3

#### BloodGen3 module enrichment analysis

Gene set variation analysis (GSVA) was performed to calculate enrichment scores for each of the 38 BloodGen3 module aggregates across all samples using normalized expression.^42,43^ GSVA enrichment scores were computed using the GSVA R package (v1.46.0) with default parameters, generating a per-sample enrichment score (from -1 to +1) for each module, where values represent the degree of coordinated down- or up-regulation of genes within each functional module.^42,43^

#### Baseline correction

To control for intra-individual baseline variation, participant-specific median enrichment scores were calculated from pre-exposure samples (timeline category "before") and subsequently subtracted from each of the following timepoint measurements corresponding to that participant. This yielded baseline-corrected enrichment scores that reflect changes relative to each participant’s pre-exposure state.

#### Group comparisons

Differential enrichment between wildfire smoke exposed and non-exposed participants was assessed using two-tailed Student’s t-tests on baseline-corrected enrichment scores during exposure period (timeline category “during”). P-values were adjusted for multiple comparisons by controlling the false discovery rate (FDR) using the Benjamini-Hochberg procedure. Bloodgen3 modules and aggregates with adjusted p-values < 0.05 were considered statistically significant.

#### Data visualization

Hierarchical clustering was performed using Ward’s method with Euclidean distance to organize samples and module aggregates based on similarity in enrichment patterns. Heatmaps were generated to visualize baseline-corrected enrichment scores, with red indicating increased enrichment and blue indicating decreased enrichment relative to baseline. Statistical significance of between-group differences was visualized using bar plots of -log10(FDR-corrected p-values), with a threshold line at p = 0.05. To contextualize exposure level, outdoor PM_2.5_ concentrations were displayed as bar plots aligned with sample collection timepoints to contextualize exposure levels.

#### Software and computational tools

All statistical analyses were performed using Python 3.9 and R 4.2.0. Data manipulation employed pandas (v1.4.2) and numpy (v1.22.3). Statistical testing and multiple comparison correction (FDR) were implemented using scipy.stats (v1.8.1) and statsmodels (v0.13.2). Data visualizations were generated using matplotlib (v3.5.2) and seaborn (v0.11.2). Complex heatmaps with hierarchical clustering were created using the ComplexHeatmap R package.

## RESULTS

### homeRNA allows for investigating the effects of wildfire smoke exposure across a large geographic area (Western and South Central U.S.)

homeRNA is a kit that allows for the self-sampling and stabilization of whole blood RNA. It consists of a commercially available blood sampling device, the Tasso-SST and a custom designed stabilization tube. Study participants were mailed homeRNA kits throughout the study, where they sampled and stabilized their own blood in response to wildfire smoke exposure. Our study design consists of three stages of homeRNA sampling: 1) collecting baseline blood samples before wildfire smoke exposure, 2) collecting exposure samples during wildfire smoke exposure, and 3) collecting post-exposure samples three and six months after wildfire smoke exposure (Figure 1). At each stage, participants were sent three homeRNA kits and asked to self-collect and stabilize blood samples every 3-4 days. After each sample collection, the participant then shipped their homeRNA-stabilized blood sample back to the study team location (Seattle, Washington).

**Figure 1.**
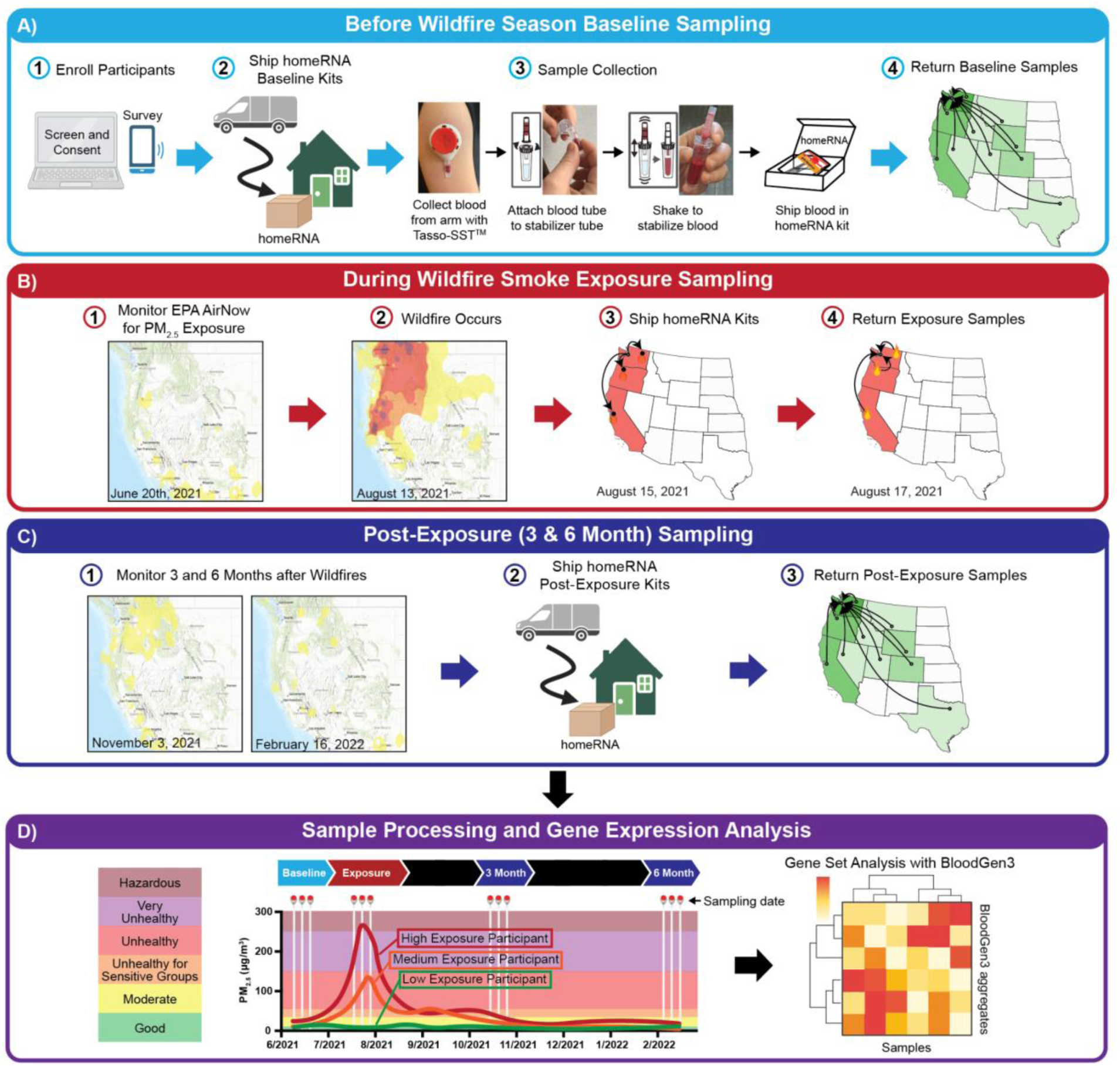
Study design for using homeRNA to investigate the effects of wildfire smoke exposure. A) Participants were enrolled across the Western and South Central United States and sent three homeRNA kits to collect baseline blood samples, which were then returned to the study team (Seattle, Washington). B) Throughout the 2021 wildfire season (June-September), wildfire occurrence and smoke exposure (PM_2.5_) were monitored using the EPA AirNow site. When a wildfire occurred, three homeRNA kits were sent to participants experiencing wildfire smoke exposure to collect exposure samples. Eighteen participants were located in Okanogan County in Washington, where two severe wildfires occurred. A subset of participants located outside of Okanogan County were also sent three homeRNA kits and sampled at similar time to serve as low exposure controls. C) All participants were sent three homeRNA kits between October-early November (∼3 months after wildfire events in Okanogan County), then again in February-April (∼6 months after wildfire events in Okanogan County) to collect post-exposure samples. D) Representative PM_2.5_ exposure timeline from a participant’s nearest outdoor EPA PM_2.5_ monitor. White vertical lines indicate blood sampling timepoints. Participants were categorized by exposure (High, Medium, or Low; see Methods). RNA from subset samples was analyzed using Nanostring gene panel coupled with BloodGen3 gene-set analysis.

In the first stage of sampling, participants were sent homeRNA kits immediately after enrollment and collected up to three baseline samples that were collected every 2-4 days prior to wildfire smoke exposure (Figure 1A). Active enrollment occurred during the beginning of June to mid-July 2021. We note that for many participants this was a true baseline, as they did not experience wildfire smoke exposure, but for some participants, particularly those located in Okanogan County who experienced smoke earlier than expected, some of the three “baseline” samples (0-2 samples of the 3 baseline samples) were collected during a transported wildfire smoke event from a fire in Canada at the end of June that caused moderate AQI due to smoke exposure in the region. Even though there was some moderate exposure during this time we still considered these as baseline samples in our analysis, as the magnitude of difference between the exposure during this time (moderate PM_2.5_ range from 9.1 - 35.4 µg/m^3^) was substantially less than the exposure these participants experienced later in July 2021 (very unhealthy PM_2.5_ 125.5 - 225.4 µg/m^3^ to hazardous PM_2.5_ ≥225.5 µg/m^3^).

In the second stage of sampling, the study team monitored the daily average PM_2.5_ level and wildfire occurrence in each participant’s location using the Environmental Protection Agency (EPA) AirNow website (https://fire.airnow.gov/). When the first wildfire was reported (located in Okanogan County), participants were immediately shipped a package containing three homeRNA kits to serve as wildfire smoke exposure samples (Figure 1B). In our particular study, a second wildfire was also reported in Okanogan County one week after the first event, so an additional package containing three homeRNA kits were sent to participants in Okanogan County. Each exposure sampling window lasted for about 1.5 weeks, thereby capturing multiple time points throughout a wildfire event (e.g., before, during, and after the wildfire-specific PM_2.5_ spike). Participants who did not experience high exposure from the Okanogan County wildfires also received three homeRNA kits at similar time points to serve as low to moderate exposure comparison groups. Stage two sampling took place during mid-July to the end of September 2021. Lastly, in the third stage of sampling, post-exposure samples were collected from all participants to assess the potential longitudinal effects of wildfire smoke exposure (Figure 1C). In this case, participants were sent a first set of three homeRNA kits three months (October to early November 2021) after the wildfire events in Okanogan County and a second set of three homeRNA kits six months (February to early April 2022) after the wildfire events. After completion of the study, each participant’s daily average PM_2.5_ exposure from wildfire smoke was recorded based on each participant’s nearest EPA PM_2.5_ monitor (see Methods for details) to assess overall exposure levels due to wildfire smoke exposure (Figure 1D).

In the present study, a total of 119 individuals were screened beginning in June 2021 (Figure 2); of these individuals screened, 10 were not eligible and 17 did not finish the screening process. Of the 92 eligible individuals, 63 participants were enrolled into the study for homeRNA sampling, with preference given to those located in Okanogan County, Washington (WA), as well as individuals who added to geographic diversity. Of the 63 enrolled participants, 5 did not collect any samples. In the end, 58 participants across 10 Western and South Central U.S. States completed the study (Washington, Oregon, California, Colorado, Idaho, Montana, Nevada, Utah, Wyoming, and Texas) and we captured two wildfire events that occurred in Okanogan County, WA (Figure 3A). We received a total of 635 homeRNA-stabilized samples before, during, and after the wildfire season. Of the 58 sampled participants, the median age was 41 (range 20 -76), 71% (n=41) reported female sex at birth, 29% (n=17) reported male sex at birth, 50% (n=29) were from Washington, and 31% (n=18) were from Okanogan County (Figure 3B).

**Figure 2.**
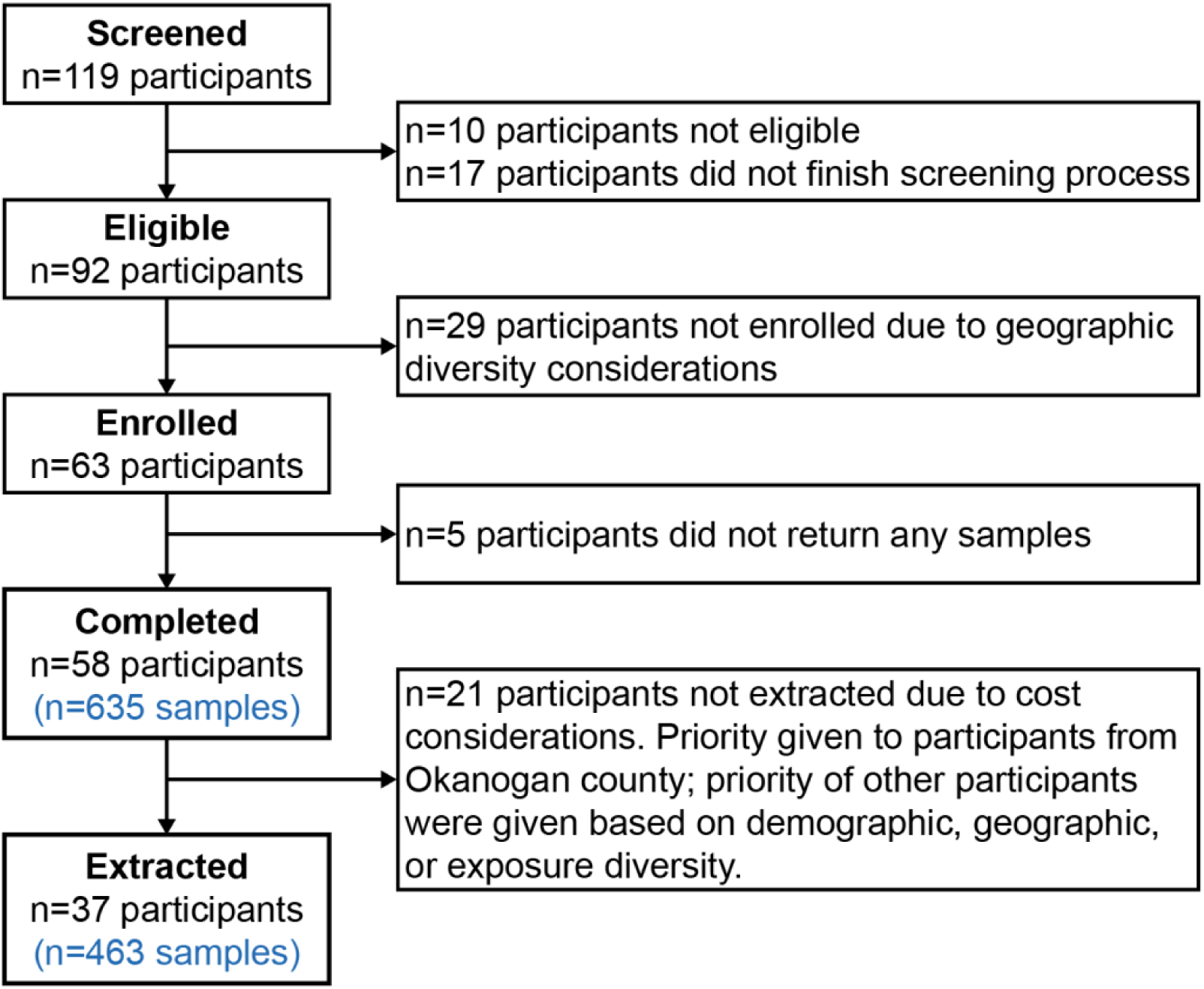
Study participant and sample flow chart. Participants were selected with a preference given to geographic diversity and those in highly wildfire-prone areas.

**Figure 3.**
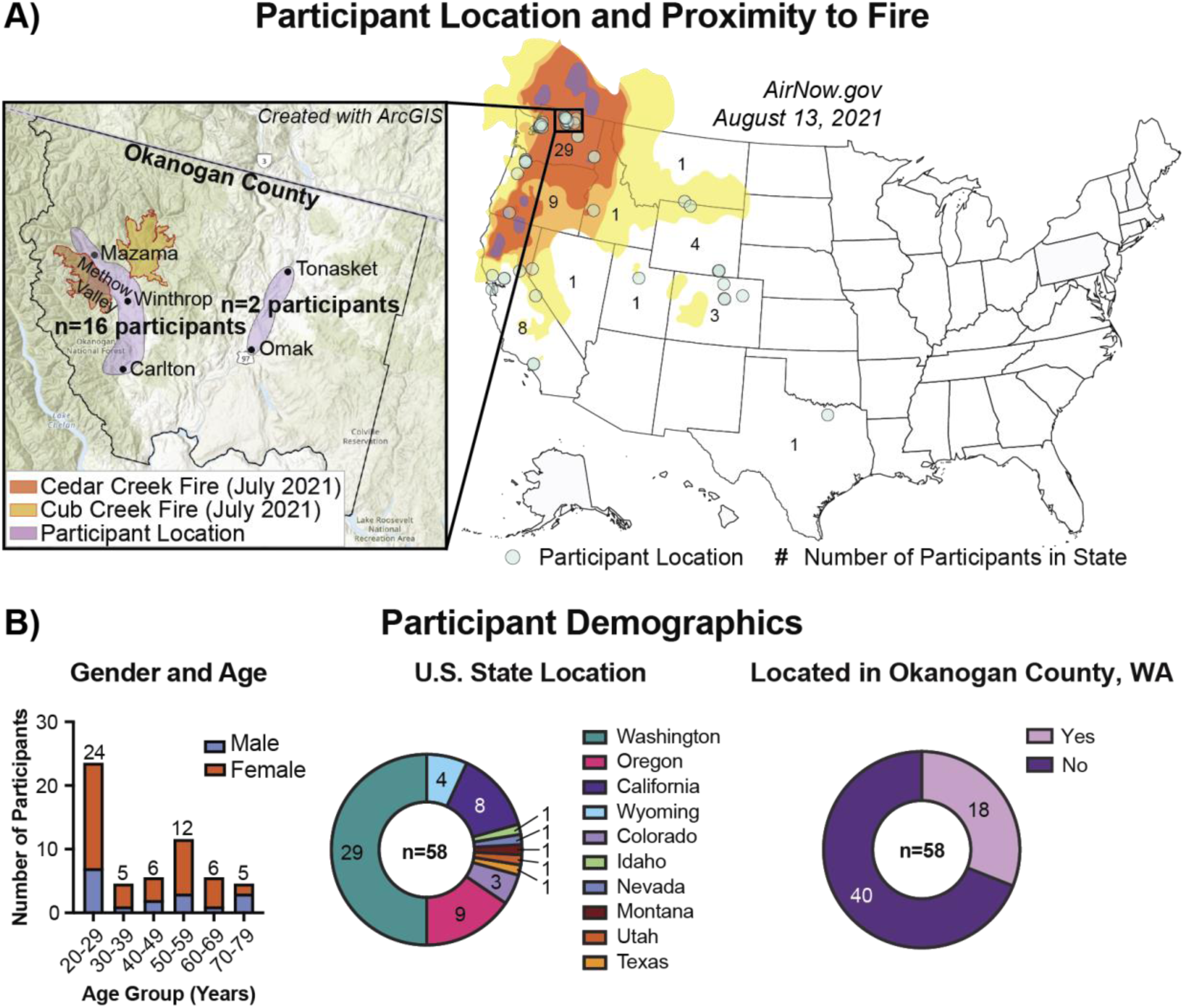
Location and demographics of study participants. A) Location of study participants in the Western and South Central United States. Each participant is represented by a circle and each state is colored based on the number of participants located within the state. PM_2.5_ levels from August 13, 2021 are shown as an overlay on the map, with colors representing the air quality index, available from the EPA AirNow interactive air quality map (airnow.gov/aqi/). Inset depicts Okanogan County in Washington state, where 18 total participants are located, with the burned area of two major wildfires, the Cedar Creek and Cub Creek fire, that started in July 2021. Inset map was generated with ArcGIS software, where each participant’s reported zip code was used to generate coordinates (arcgis.com). Fire boundaries were provided by Central Washington Fire Recovery (centralwashingtonfirerecovery.info/wildfire-reports/). B) Demographics of all sampled participants who completed the study (n=58 participants), including gender, age, US state location, and number of participants located in Okanogan County, which was the area of the two major wildfires captured in this study.

#### homeRNA is an-easy-to use kit that allows high participant retention and flexibility across a 10- month longitudinal study

Spanning 10 months (June 2021-April 2022), each participant collected up to 15 blood samples with the homeRNA kit before, during, and after the wildfire season; an average of 11 samples were self-collected from each participant. Despite the extended study duration and frequent sampling requirements, we achieved remarkably high study retention. Of the 58 participants who completed at least one sample at the start of the study, 93% (n=54/58) collected at least 6 samples. All of the study participants were also asked to sample 3-months post wildfire smoke exposure (between October and November 2021), and 95% (n=55/58) completed at least one of the three requested samples (samples were scheduled to be every 2-4 days for each set of 3 samples). For the 6-month post exposure samples (between February to April 2022), 37 participants were asked to complete sampling and 95% (n=35/37) completed at least one of the three requested samples; this was at the end of 10 months total time after initial enrollment in June 2021 in the study. In the study conclusion survey, 93% of participants (n=50/54 participants who completed the closing survey) expressed interest in participating in a second year of the study during the 2022 wildfire season.

To assess the usability and practicality of using homeRNA in a 10-month longitudinal study, each participant completed a sample collection survey (see methods for details) that included questions regarding the usability of the homeRNA kit each time they sampled. As participants collected up to 15 samples in this study, we were first interested in participants’ perceptions the first time they used the kit. After collecting their first blood sample, the majority of participants took a total of 5-10 minutes to sample and stabilize their blood with homeRNA and reported that the Tasso-SST device and the custom-engineered stabilizer tube were easy to use (Figure 4A). Additionally, all participants reported no or mild pain (n=35/58 reported no pain and n=23/58 reported mild pain) associated with the blood collection using homeRNA for the first time (Figure 4A). These initial responses suggest that the homeRNA kit was easy to use even during the first attempt.

**Figure 4.**
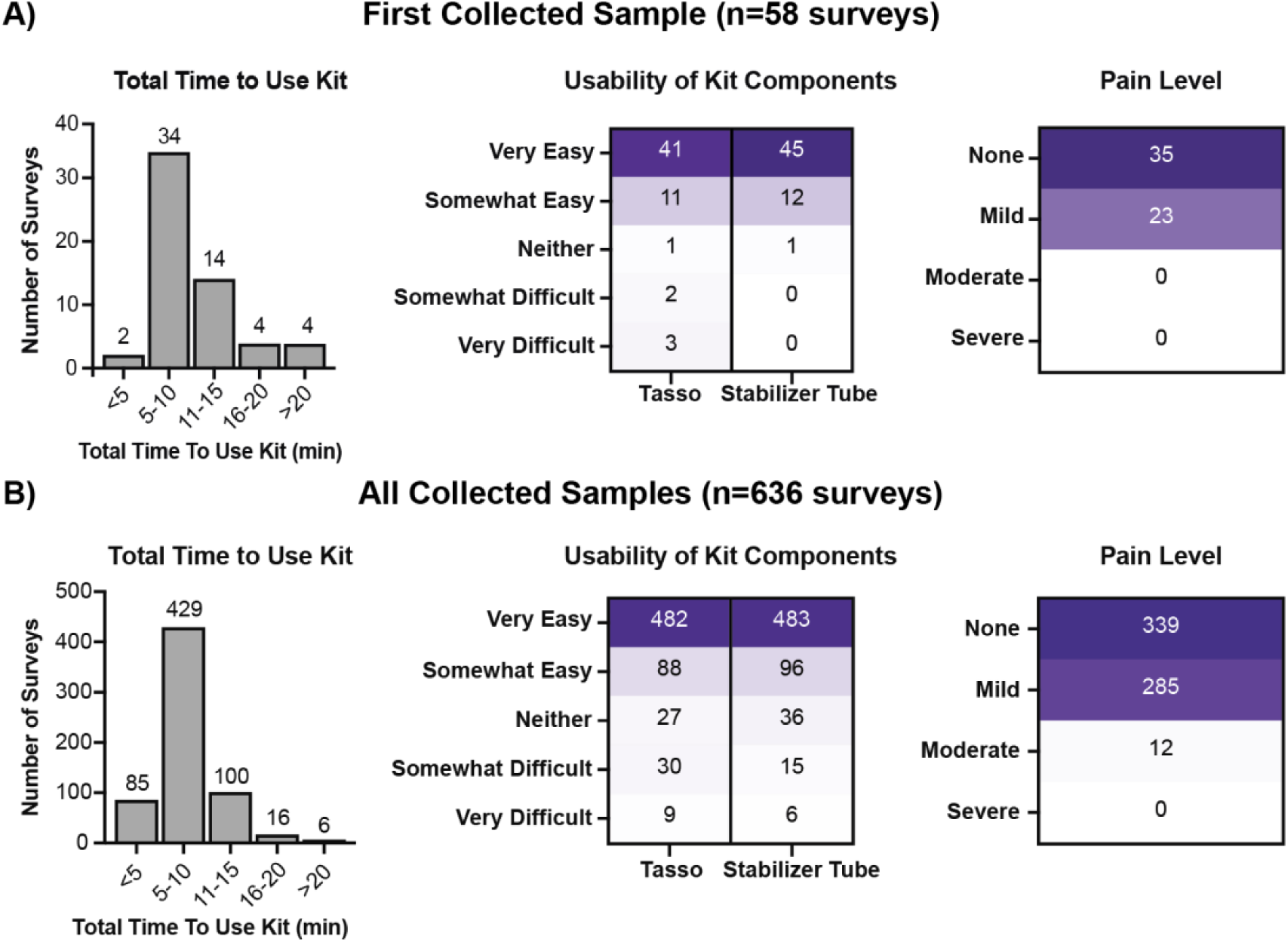
Usability survey responses for using the homeRNA kit to self-collect and stabilize blood throughout the study. Participant survey responses on the total time to use the homeRNA kit (left), the usability of the Tasso-SST blood collection device and the stabilizer tube (middle), and the pain level associated with blood collection (right) from A) first collected sample (n=58 surveys) and B) all collected samples (n=636 surveys) from 58 participants.

We collected a total of 636 sampling surveys throughout the study. We note that while we collected 636 surveys, we received 635 homeRNA-stabilized blood samples; notably, throughout our 10-month study, only one sample was missing. When all the sampling surveys were assessed, the questions related to the homeRNA kit’s usability showed similar responses as their first survey response (Figure 4B) The similar survey responses between the first sample collection and all sample collections demonstrate that homeRNA was easy to use in the beginning and stayed easy to use throughout the study. Overall, the survey responses regarding the usability of homeRNA suggest the effectiveness of deploying homeRNA in a remote study design.

Apart from the kit being easy to use, the flexibility of the study design enabled by the remote self-blood sampling and stabilization further increased user’s perception and willingness to be included in a study using homeRNA. A follow-up survey was administered in November 2023, 18 months after the completion of the study on the 2021 wildfire season, to assess participants’ preference for the remote study design over a clinic-based blood draw study design and overall perception of a homeRNA-based remote study. In this survey, 93% of the respondents (n=28/30) considered participating in a remote blood sampling study (the current study) to be “significantly easier” or “somewhat easier” than an in-person blood sampling study (e.g., clinic-based blood draw) (Figure 5A), reporting that length of commute to a clinic and difficulty fitting a clinic visit into their schedule was a barrier to in-person participation (Figure 5D). In contrast, 77% (n=23/30) reported that the flexibility in sampling time was an important factor in remote-sampling participation (Figure 5B). Given that this study design could be applied to investigate transcriptomic responses during various natural hazards (e.g., wildfires, earthquakes, extreme weather), we assessed participant comfort with using homeRNA kits during emergency situations. Notably, 83% (n=25/30) of respondents reported that they would be “very comfortable” or "somewhat comfortable” with using homeRNA during a disaster resulting from a natural hazard (Figure 5C). Lastly, almost all respondents (n=29/30) indicated that they would be willing to participate in this or a similar remote blood sampling study again (Figure 5E), with 70% (n=21/30) reporting that they would participate in a similar remote blood sampling study for up to 5 years (with this option being the longest period we provided in the survey) (Figure 5F).

**Figure 5.**
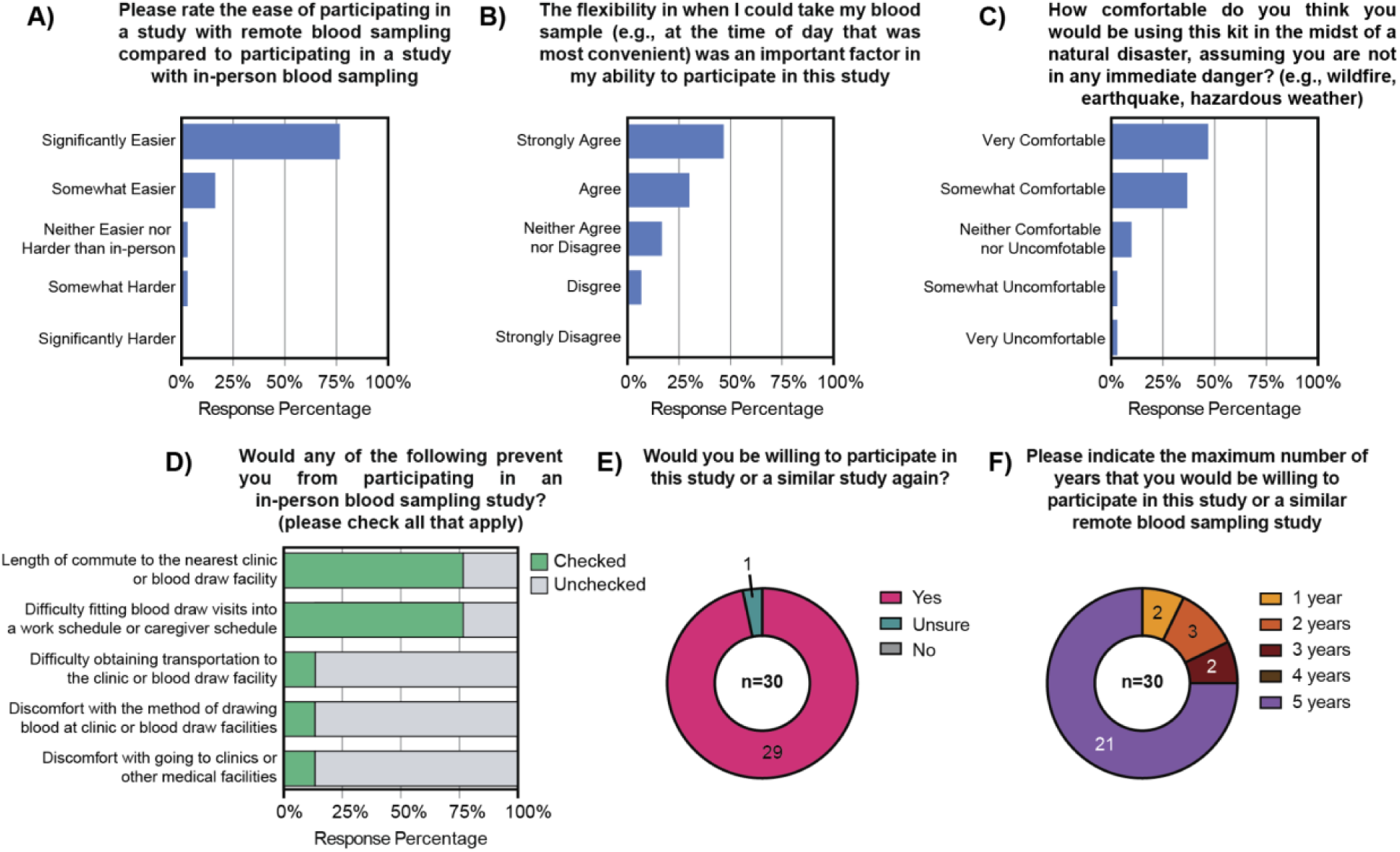
Follow-up survey responses from 30 participants (out of 40 participants invited) who were surveyed 18 months after the completion of the 10-month long wildfire smoke exposure study (June 2021-April 2022). A) Survey response on the A) ease of participating in a remote blood sampling study compared to an in-person blood sampling study, B) flexibility provided by remote-based sampling, and C) comfortability of homeRNA-based sampling during a disaster. D) Checklist response on barriers to participating in an in-person blood sampling study. E) Survey response on willingness to participate in a similar remote blood sampling study again and F) maximum number of years willing to participate in a similar study again. Full survey questions are available in the Supplementary Information.

#### Our flexible study design captured a wide range of PM_2.5_ exposure throughout wildfire season

Our study captured two major wildfires, both of which occurred in Okanogan County, WA in July 2021: the Cedar Creek Fire and the Cub Creek Fire (Figure 3A). Therefore, we will discuss the 58 sampled participants in the context of three groups based on their daily average PM_2.5_ exposure: 1) high exposure participants (n=20 participants), which were 17 participants located in Okanogan County who were exposed to active wildfire smoke and three participants located in Nevada, Oregon, and California who were exposed to active wildfire smoke from the Caldor fire in Northern California and Nevada, 2) medium exposure participants (n=5 participants), which were participants who had elevated PM_2.5_ levels in the unhealthy AQI category due to indirect wildfire smoke exposure (one participant was in Okanogan County farther away from the active wildfire, three participants were in Washington outside of Okanogan County, and one participant was in California who experienced a wildfire event later in the study), and 3) low exposure participants (n=32 participants) located in all other sampled states/counties who were not exposed to wildfire smoke and experienced good or moderate AQI categories based on our internal cut-off for how we defined PM_2.5_ exposure (see extended details in Methods). The daily average PM_2.5_ data (indoor and outdoor) of an example participant from each exposure group across the study timeline (June 2021 - April 2022) was plotted against their homeRNA sampling dates (Figure 6A); these graphs are color coded based on the EPA’s AQI categories based on PM_2.5_ exposures (Table S1). A subset of participants were selected to receive Indoor PurpleAir sensors to track indoor PM_2.5_ concentrations. The daily average indoor Purple Air sensor data is plotted in grey on Figure 6A for three example participants, along with the outdoor EPA PM_2.5_ monitors in black. The outdoor EPA PM_2.5_ monitor data for the nine participants with samples analyzed with the Nanostring gene panel are plotted in Figure S3, with indoor PurpleAir PM_2.5_ also plotted when available.

**Figure 6.**
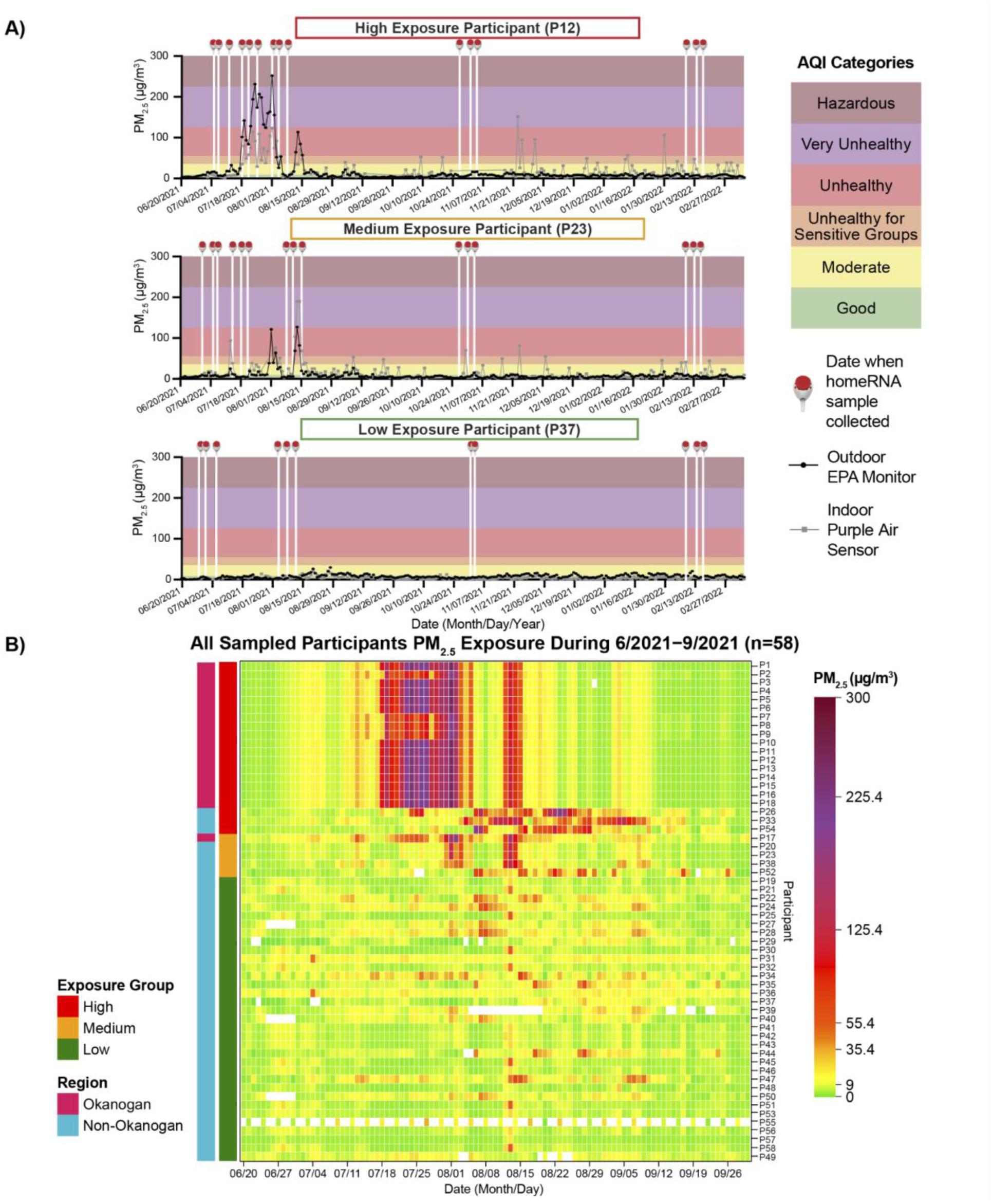
homeRNA was able to capture a wide range of participant PM_2.5_ exposures across the study period. A) Data from example participants exposed to high, medium, or low levels of wildfire smoke, plotted against the homeRNA sampling timepoints (white vertical lines with Tasso symbols on top), outdoor EPA PM_2.5_ monitor (black lines with data points), indoor PurpleAir sensor (grey lines with data points), and AQI categories (colored background). B) Heatmap summarizing the outdoor PM_2.5_ levels during the wildfire season (June 2021-September 2021) for all 58 sampled participants. Heatmap rows represent individual participants and columns represent daily average PM_2.5_ concentrations for each participant throughout the 2021 wildfire season, with each PM_2.5_ cell colored by the corresponding AQI category. Cells colored white represent days where no PM_2.5_ concentrations were collected by the participant’s nearest EPA PM_2.5_ monitor. Participants are color-coded based on their geographic region (magenta: Okanogan County, teal: Non-Okanogan County) and exposure levels (red: high exposure, orange: medium exposure, green: low exposure).

To visualize the PM_2.5_ exposure for all participants throughout the wildfire season, we summarized the outdoor PM_2.5_ data of all 58 sampled participants during June through September 2021 in a color coded heatmap based on the EPA’s AQI categories (Figure 6B). In this heatmap, we grouped participants based on their wildfire exposure (high, medium, or low) to visualize the differences in PM_2.5_ ranges captured in our study. This visualization helps illustrate temporal patterns in PM_2.5_ exposure across the sampled cohort, highlighting the periods of elevated PM_2.5_ levels during the Okanogan County Cedar Creek and Cub Creek wildfires in late July and early August 2021. The sporadic elevated PM_2.5_ levels visible in the medium exposure group throughout August and September demonstrate the variable nature of indirect wildfire smoke exposure. White squares in the heatmap represent days where PM_2.5_ concentrations were not reported by the EPA PM_2.5_ monitor for that participant’s location, which constitutes a potential limitation of this study as it may result in incomplete exposure characterization for some participants during certain time periods.

#### homeRNA demonstrated sufficient RNA quality throughout study

At the end of the study, a total of 635 homeRNA samples were returned from 58 participants. Total blood cellular RNA was extracted from 463 samples for quality assessment. RNA integrity number (RIN) values were assessed across all participant samples to evaluate RNA quality from stabilized blood collections. Of the extracted samples, the average RIN value was 7.4 ± 1.3 with 92% of samples (n=426/463) above a RIN value of 6 (Figure 7A), which was the cutoff RIN value we used for downstream transcriptomic analysis; the 455 samples for which a RIN value was obtained are reflected in Fig 7A (eight samples did not yield a RIN value). Despite the presence of some degraded samples, the majority of samples yielding RIN values ≥ 6 indicates successful RNA preservation in our study, which is in concordance with our previous literature.^58–62^ Collection efficiency was also high, with most participants collecting blood samples within 3-5 minutes and most samples had at least 300 μL of blood (n=412/636 surveys reported blood level 3 or 4) (Figure S1). Additionally, the distribution of RIN values varied both within and between participants, with low RIN values (< 6) occurring sporadically across the cohort rather than clustering within specific individuals, and participant age was also not predictive of RNA quality (Figure S2).

**Figure 7.**
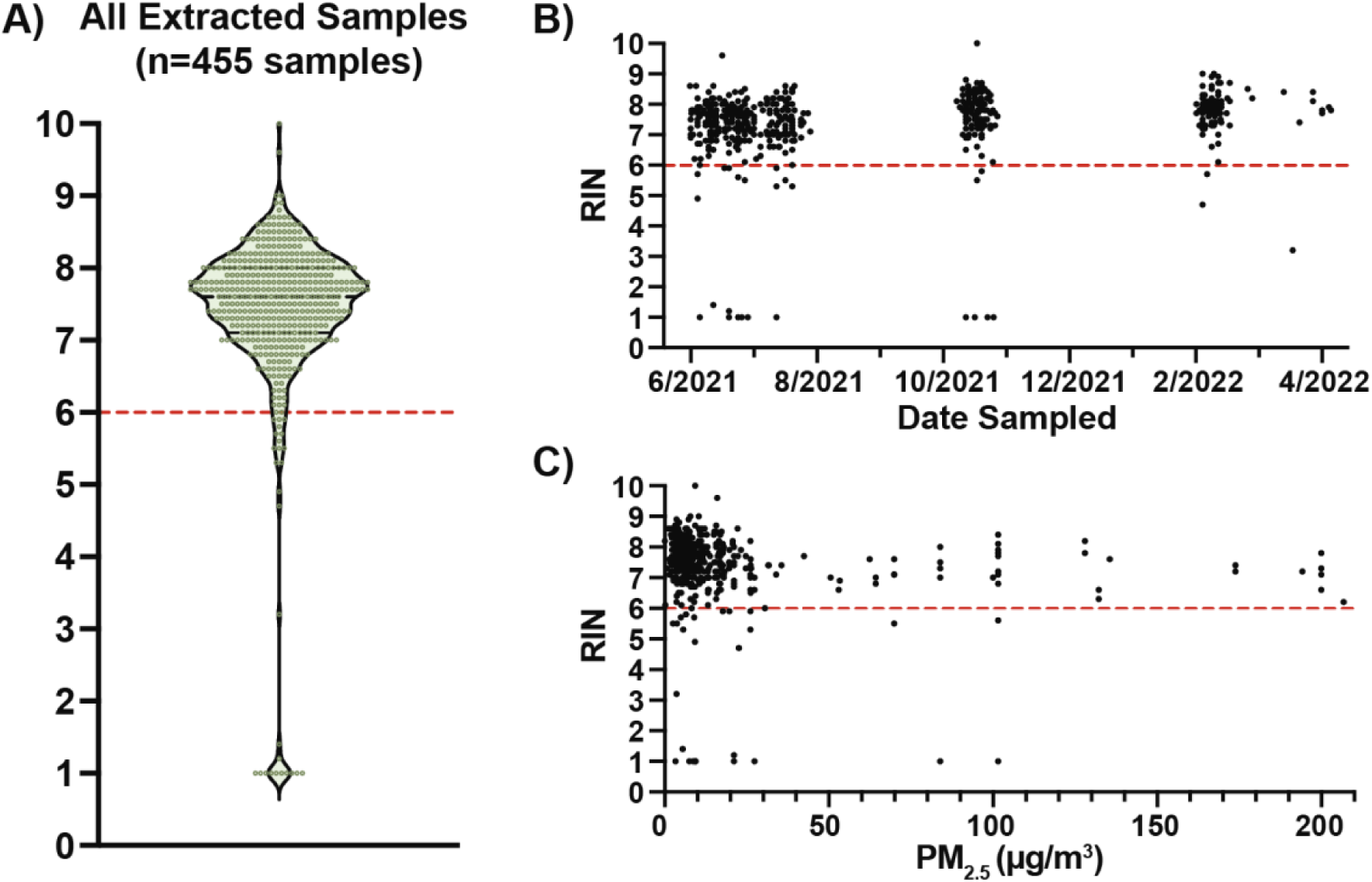
homeRNA was able to sufficiently stabilize RNA and preserve integrity throughout the 10-month long study regardless of sampling season and participant exposure. A) Resulting RNA Integrity Number (RIN) values of all extracted samples (n=455 samples). Eight samples did not yield a measurable RIN value. RIN values are given on a scale from 1 to 10, with a RIN of 1 representing the most degraded RNA and a RIN of 10 representing the most intact RNA. We used an internal quality control cutoff at RIN = 6 for downstream transcriptomic analysis. B) RIN values according to the date of sample collection and stabilization throughout the study period (June 2021 – April 2022). C) RIN values according to the corresponding average PM_2.5_ exposure on the date of sampling.

To assess potential confounding factors in our home-based sample collection protocol, we examined RIN values in relation to sampling date and PM_2.5_ exposure levels. Sample collection occurred over a 10-month period and RIN values were consistently adequate throughout the sampling timeline, suggesting that seasonal variations did not systematically affect sample integrity (Figure 7B). Similarly, PM_2.5_ exposure levels, which varied across participants due to wildfire smoke events, did not negatively affect RNA stabilization and subsequent RIN values (Figure 7C). These findings demonstrate that our study successfully captured high-quality RNA samples across diverse temporal and environmental conditions, suggesting that homeRNA was robust to external factors that might otherwise introduce bias into RNA quality assessments. This further reinforces our previous studies where we found minimal effects of shipping time and temperature on homeRNA-stabilized samples.^38,39^

#### Gene expression analysis of subset of participants shows preliminary patterns suggesting inflammatory changes with gene set analysis

In a preliminary analysis, we analyzed 107 samples from nine participants (out of 463 extracted samples from 37 participants) using a targeted Nanostring Autoimmune gene panel. The Autoimmune panel includes 770 genes that encompass 35 pathways and processes associated with autoimmune disease and chronic inflammatory disorders; in particular, we were interested in capturing a wide array of inflammation markers, based on previous studies implicating an inflammatory-like response to wildfire smoke exposure.^21,28,29,31,36^ Since there is no established transcriptional marker for wildfire smoke exposure, we chose to map the Nanostring gene expression data to the BloodGen3 repertoire framework, which contextualizes inflammatory signatures within broader immunological networks, rather than examining individual genes in isolation. BloodGen3 is a fixed repertoire of 382 transcriptional modules (which cluster into 38 module aggregates) derived from gene co-expression patterns across 985 blood samples from 16 diverse immunological and physiological states (including bacterial and viral infections, autoimmune diseases, COPD, cancer, and pregnancy).^42,43^ This fixed repertoire provides a stable framework for analyzing and interpreting blood transcriptome data by grouping co-regulated genes into functionally annotated modules. Beyond the 16 cohorts used to construct the modules, this repertoire has since been applied to multiple independent datasets spanning diverse conditions and physiological states, providing a consistent reference for interpreting blood transcriptional profiling data and enabling cross-study comparisons.^44–49^

The nine participants for the preliminary analysis were chosen to include both participants from Okanogan County primarily (n=6, P5, P6, P8, P10, P12, P17), along with some participants with low exposure located in California (n=2, P31, P37) and medium exposure located in central Washington (n=1, P23), which experienced moderate smoke exposure from the Okanogan County fires. First, we performed a baseline correction on each participant’s exposure timepoint samples by subtracting each participant’s median gene module enrichment score calculated from their baseline timepoint samples. This correction allows for each participant to serve as their own control, thereby allowing us to identify changes in gene module activity relative to each participant’s individual pre-exposure (i.e., baseline) state, this improves sensitivity for detecting exposure-related changes, while minimizing confounding effects of baseline differences We then compared the baseline-corrected enrichment scores between high-smoke-exposed participants (from Okanogan County) and low-medium-exposed participants during the active wildfire period to identify modules that responded differently to smoke exposure. Of note, “exposure” timepoint samples from the low and medium smoke exposure participants (P23, P31, and P37) represent the time that they were collected, not the level of smoke exposure; these participants were sampled at the same time as the participants in Okanogan County who were the only group to actively experience wildfire smoke in the context of this analysis.

At the individual gene level, we identified 253 genes with differential expression (FDR < 0.05) between exposed and non-exposed groups. To determine biological significance, we also performed a minimum-effect test where the null hypothesis was |log₂FC| ≤ 0.5.^50^ No genes achieved FDR < 0.05 using this more stringent approach, indicating that while statistical differences exist, no genes showed fold changes greater than 1.4-fold with controlled false discovery rate. However, small transcriptional changes can still reflect biologically meaningful differences, particularly for regulatory genes and transcription factors that may not elicit large expression changes. Given our smaller panel (∼770 genes) and limited sample size (n=6 vs 3), we present these findings as preliminary observations requiring further validation.

We next evaluated transcriptomic changes at the gene set level using the BloodGen3 framework. Of the 770 genes present in the Nanostring panel (20 of which are housekeeping genes), 567 genes were present in the BloodGen3 framework comprising 382 modules. These 382 modules are further organized into 38 higher-order module aggregates based on functional and expression similarities, providing both granular (module-level) and broader (aggregate-level) perspectives on immune system activity. At the module level, we evaluated enrichment across all 382 modules that comprise the BloodGen3 framework. We observed 10 modules representing distinct immune functions with differential expression patterns between the high-exposure wildfire exposed participants (i.e., Okanogan County participants, n=6) and the non-exposed participants (i.e., participants in other locations, n=3) (FDR < 0.05) (Figure 8A). These included modules associated with inflammation (M13.1 and M13.12 overexpressed in high exposure group), B cells (M13.18 underexpressed in high exposure group), T cells (M16.24 and M15.38 underexpressed in high exposure group), prostanoids (M8.2 overexpressed in high exposure group), and monocytes (M15.7 underexpressed in high exposure group). The overexpression of the prostanoid module (M8.2) in the high exposure group suggests activation of cyclooxygenase-derived lipid mediators, which have been found to promote inflammation while suppressing adaptive immunity through prostaglandin E2 (PGE2) signaling^51^. The underexpression of the monocyte module (M15.7) in the high exposure group could potentially reflect monocyte redistribution rather than true suppression, as previous studies have demonstrated that particulate matter exposure can stimulate bone marrow release of monocytes with subsequent migration to lung tissue^52,53^. At the BloodGen3 aggregate level, we examined the 38 higher-order module aggregates that represent major immunological and physiological processes (Figure 8B). Of the 38 aggregates, we observed eight aggregates with differential expression patterns in the wildfire exposed participants compared to the non-exposed participants. Four of these aggregates did not have annotated functions. The other four aggregates had annotation functions of lymphocytic (aggregates A1 and A6, both underexpressed, suggesting potentially reduced adaptive immune cell activity or other mechanisms) and inflammation (aggregates A33 and A35, both overexpressed, possibly indicating increased innate immune activation and inflammatory mediator expression); in previous work, a similar expression pattern was found to be associated with psoriasis.^49^ This pattern of decreased circulating lymphocyte-associated transcripts alongside increased inflammatory markers is consistent with complex stress-induced immune modulation, potentially involving lymphocyte redistribution and inflammatory activation^43,54^. Further, the presence of altered but functionally unannotated modules and aggregates is expected within the BloodGen3 framework, as modules are defined purely by co-expression patterns across diverse immunological states rather than by predetermined functional categories^43^. These unannotated modules and aggregates may represent novel coordinated transcriptional responses specific to environmental exposures like wildfire smoke or biological processes not yet well-characterized. Their alteration suggests wildfire smoke exposure may affect transcriptional networks beyond classical immune pathways, which warrants future investigation to determine their biological significance.

**Figure 8.**
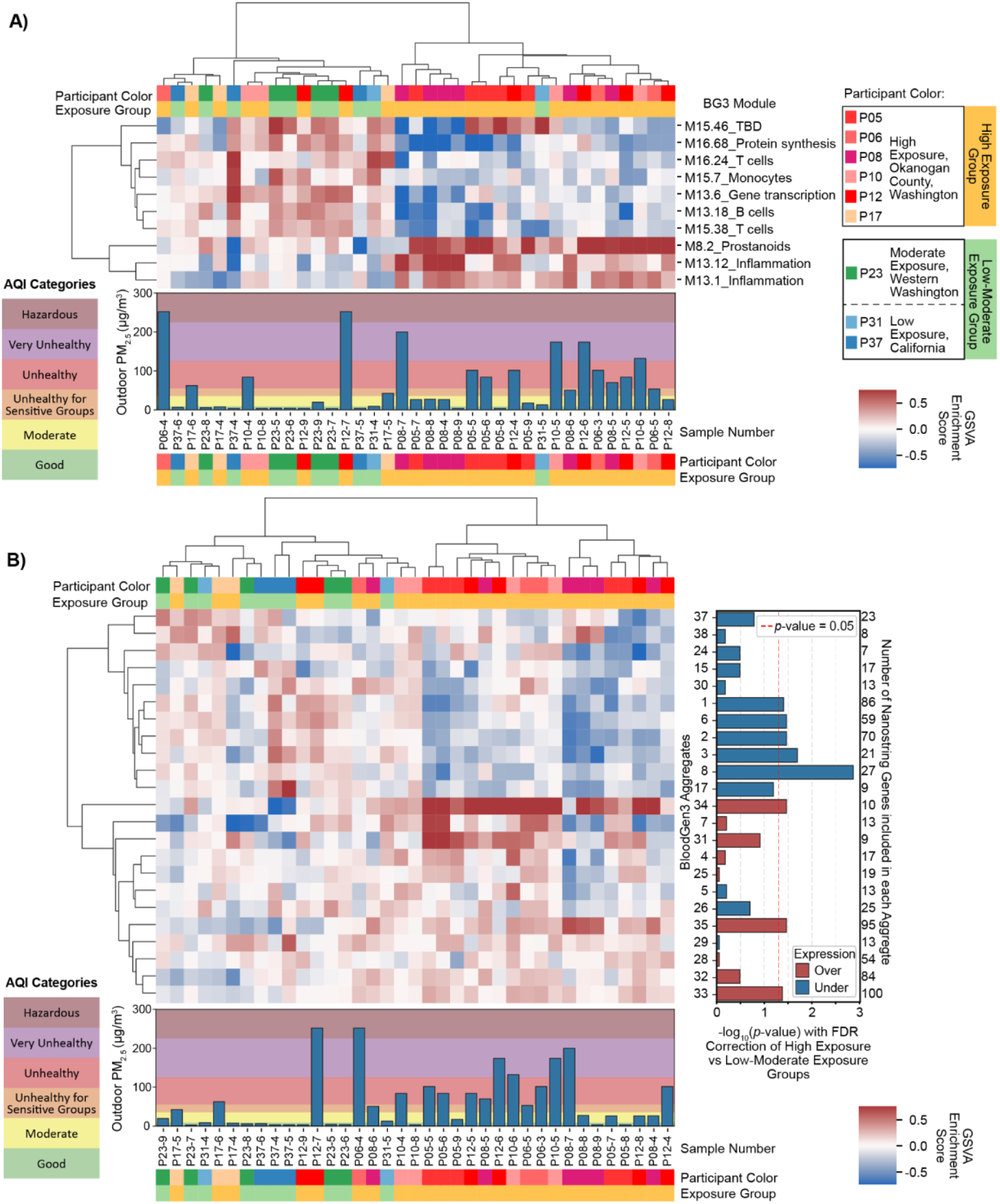
Heatmap of wildfire smoke exposure gene expression using BloodGen3 framework. Blood samples from nine participants (six high-exposure from Okanogan County, three low-moderate exposure from other locations; n=107 samples total) were analyzed using the Nanostring Autoimmune panel (770 genes), with 567 genes mapped to the BloodGen3 framework. A) Module-level and B) aggregate-level heatmaps show gene set variation analysis (GSVA) enrichment scores after baseline correction (participant-specific median enrichment scores were subtracted from pre-exposure samples). Positive scores (red) indicate higher than average gene expression in a given module/aggregate; negative scores (blue) indicate lower than average gene expression in a given module/aggregate. Rows represent BloodGen3 A) modules or B) aggregates with hierarchical clustering; columns represent individual samples with hierarchical clustering. Sample labels indicate participant and sample number (e.g., P23 -9 represents sample 9 from participant 23). Bar plots below heatmaps show average outdoor PM_2.5_ concentrations from nearest EPA monitor on sampling dates, colored by EPA AQI categories. In B), statistical significance was assessed using two-sample Welch’s t-tests comparing high vs. low-moderate exposure groups with Benjamini-Hochberg correction for multiple testing (FDR < 0.05). Significant aggregates are shown with -log_10_(p-values) on the right; red bars indicate overexpression, blue bars indicate underexpression.

## DISCUSSION

This study demonstrates that homeRNA technology enables flexible remote study logistics for investigating gene expression changes due to wildfire smoke exposure with geographically distributed participant recruitment. With homeRNA, participants can easily self-collect and stabilize blood samples in their homes throughout extended study periods, overcoming the inherent challenges of long-term, in-person blood collection studies. In this study, we employed a dual recruitment strategy: broad enrollment across the Western and South Central U.S., combined with targeted recruitment in Okanogan County, Washington, a region with historically severe wildfire seasons and active community wildfire preparedness efforts. This approach both allowed us to capture participants at similar time points who were either exposed or not exposed to wildfire smoke and to increase the likelihood of capturing a wildfire event in a subset of participants.

The utilization of a remote study design with homeRNA allowed for the study team to react to wildfire smoke exposure in real time, often sending out study kits within a day of the onset of a wildfire smoke event. Such flexibility would not have been possible in a more rigid study design requiring a physical location for blood draws. Moreover, once in participants hands, the homeRNA sampling could travel with the participants in the case of an evacuation. In fact, at one point in the study, some participants needed to evacuate, and a subset of these participants chose to take their homeRNA kits with them and continued sampling; such anecdotes exemplify the flexible nature of homeRNA study design and the potential utility of such tools for the study of disasters broadly. In this case, the study team used their evacuation address to assess their exposures and to schedule a UPS pick up of their samples for those days. Notably, despite the psychological stress that wildfires and other disasters can impose on affected populations, the majority of surveyed participants reported being very or somewhat comfortable using homeRNA during such events, suggesting using the homeRNA kit does not cause additional burden during already challenging circumstances.

Further, it was easier for study participants to collect blood samples, both from the participant perspective (i.e., not needing to go to a physical location to have a blood draw) and also from a study logistics perspective (i.e., not needing to set up and staff a physical clinic or phlebotomy site for participants). These factors make collecting more samples in a longitudinal remote study simpler and less costly; thus, collecting up to 15 blood samples across 10 months with high participant retention and sample return rate to the last sampling time point is not only feasible, but demonstrated with this study. Additionally, the ability to collect baseline or pre-exposure samples allows for every participant to serve as their own control.

This study provides a rare window into real-world gene expression responses during an active wildfire season, with participants self-sampling throughout actual smoke exposure events in their home environments. By the nature of the remote study design, we sampled from the general population while they were being exposed to real-life smoke exposure in their own homes. Importantly, this includes the psychological stress inherent to experiencing a disaster, which is an inseparable component of wildfire smoke exposure. Some participants even needed to evacuate their homes during our study; such circumstances, including the associated stress, disrupted sleep patterns, and altered daily activities, are integral parts of how populations actually experience wildfire smoke events. Therefore, we cannot definitively determine whether the observed gene expression changes stem from the direct biological effects of PM_2.5_ and other smoke components, or from the stress of experiencing a wildfire event. This inability to isolate specific causal mechanisms is inherent to studying disasters as they occur. However, understanding this holistic response may be more relevant for public health purposes as communities experiencing wildfire smoke, or other disasters, will often experience these stressors.

Beyond these inherent challenges of researching disasters, our study faces additional methodological limitations in quantifying exposure and controlling for temporal confounders, particularly in how we collected daily PM_2.5_ averages. The distance between each participant’s location and their nearest EPA PM_2.5_ monitor varies amongst the study participants, with many participants, particularly those in Okanogan County, sharing the same monitor despite potentially different actual exposures. To look at differences in outdoor versus indoor PM_2.5_ exposures, we also sent a subset of participants Purple Air sensors as part of the Clean Air Methow program. We observed discrepancies between indoor and outdoor PM_2.5_ levels, with some participants experiencing higher indoor PM_2.5_ at times possibly due to other indoor environmental sources such as cooking, candles, or differences in home ventilation; participants were asked to place the PurpleAir sensors away from indoor stoves, but the ventilation and layout of homes can vary. Individual behaviors further affect effective exposure assessment depending on time spent indoors versus outdoors and protective measures such as wearing N95 masks or using HEPA filters. For the purposes of this initial analysis, we chose to not include the indoor PM2.5 levels collected by the PurpleAir sensors due to incomplete coverage across all participants and the additional complexity of modeling indoor exposure, which we plan to address in future analyses with subset comparisons. Additionally, since our study lasted 10 months, there are other potentially confounding immune response factors such as the seasonal flu and allergies, which fluctuate in severity across different seasons. While all of these combined factors are inherent in most human subject based research, they are important confounding variables to consider when designing a remote study with homeRNA and interpreting the transcriptomic results. These challenges with using proximal outdoor monitors as surrogate exposures may be overcome with the recent advent of more affordable, wearable personal exposure monitors for PM_2.5_ (e.g., Atmotube) or other emerging wildfire-specific PM_2.5_ exposure modeling frameworks.^55,56^

In the 2021 wildfire season, Okanogan County experienced one of its worst summers for wildfire smoke; smoke from nearby wildfires caused several days of hazardous AQI category (hazardous air quality is a PM_2.5_ ≥ 225.5 µg/m^3^; for reference, good air quality is PM_2.5_ ≤ 9.0 µg/m^3^). As such, the participants located in Okanogan County served as our "high exposure" group. Concurrently sampled participants enrolled elsewhere that did not experience wildfire smoke were categorized into "low exposure", and participants that experienced transported smoke exposure from the Okanogan County fires were categorized into “moderate exposure” groups. Due to cost, this initial analysis was small (n=107 samples), with only six participants experiencing significant wildfire smoke exposure and three participants experiencing low-moderate levels of PM_2.5_ exposure; additional analysis on over 300 samples from 29 participants is forthcoming.

The demographic composition of our analyzed cohort and broader study population reflects both the strengths and limitations of our remote recruitment approach. Our participants were predominantly female (71%) and had a median age of 41 years. While we attempted to match controls for age and sex when selecting the three participants with low and moderate exposure for our initial transcriptomic analysis, this matching was constrained by the limited control participant pool. Comprehensive demographic data on race/ethnicity was not collected in initial enrollment surveys, a limitation we addressed in follow-up surveys but that prevented our enrollment process from being responsive to balancing demographics. Our recruitment through word-of-mouth and existing community networks, particularly the Clean Air Ambassador Program led by Clean Air Methow, a community-based organization in Okanogan County, WA, likely contributed to demographic homogeneity, with overrepresentation of groups already engaged in community air quality initiatives. These demographic limitations should be considered when interpreting results, as wildfire smoke exposure impacts may vary across different demographic groups.

Despite the small sample size, our preliminary transcriptomic analysis in nine participants (n=6 exposed, n=3 non-exposed) identified patterns of inflammatory activation characterized by overexpression of inflammatory module aggregates (A33, A35) and inflammation-associated modules (M13.1 and M13.12). Concurrently, we observed reduced adaptive immune cell activity characterized by underexpression of lymphocytic aggregates (A1, A6), modules associated with T cells (M16.24 and M15.38), modules associated with B cells (M13.18), and monocyte modules (M15.7). While our sample size limits definitive conclusions, these initial observations using the BloodGen3 framework demonstrate its sensitivity in detecting coordinated transcriptional responses to environmental exposures. It is noteworthy that these patterns involve modules relevant to immune competence and vaccine responses. Interestingly, a recent study by Sanghar et al. documented compromised COVID-19 vaccine immunity in individuals exposed to wildfire smoke.^24^ Although we cannot directly link our preliminary observations to their clinical findings, the involvement of similar adaptive immune pathways in both studies warrants further investigation with larger cohorts to understand potential mechanisms underlying immune alterations during environmental exposures. These preliminary findings, while requiring validation in our full cohort and independent studies, suggest that homeRNA technology can overcome logistical challenges of disaster research while potentially capturing biologically relevant gene expression signatures.

Currently, we are employing bulk RNA-sequencing on over 300 samples from 29 participants from this study to further investigate wildfire-related transcriptomic response. This follow up analysis will have greater statistical power and will include a greater number of genes than were included in the Nanostring panel, strengthening the aggregate analysis. With this larger data set, we will also be able to employ additional investigations into the temporal component of the sampling data with time course gene set analysis, utilizing the 3- and 6-month timepoint samples to further understand the time course of the gene expression response to wildfire smoke.

The demonstrated feasibility and flexibility of homeRNA technology in this study opens new avenues for population-scale research on environmental exposures and natural disasters. The remote nature of homeRNA sampling addresses key barriers that have traditionally limited disaster-related health research: the ability to deploy studies quickly, maintain geographic flexibility, and preserve study continuity when communities experience disruption. Future applications could extend beyond wildfire smoke to investigate gene expression changes during other environmental exposures such as extreme weather events or infectious disease outbreaks, where traditional clinic-based sampling would be logistically challenging. Furthermore, the preliminary transcriptomic findings from this study could inform the development of more targeted, cost-effective gene panels specifically designed for wildfire smoke exposure research, enabling larger studies with greater statistical power at reduced costs. The scalability and flexibility of remote sampling technologies like homeRNA represent valuable tools for advancing our understanding of how populations respond biologically to environmental exposures, ultimately informing public health research and intervention strategies across diverse geographic regions and populations.

## Data Availability

All de-identified gene counts data and other exposure and usability data produced in the study are available upon reasonable request to the authors.

## ACKNOWLEDGEMENTS

This publication was supported by Schmidt Sciences, LLC (for bioinformatic data analysis pipeline development), the David and Lucile Packard Foundation, a STEP grant from University of Washington CoMotion, and the National Institutes of Health (NIH) through the National Institute of Environmental Health Sciences award number R21ES034338, the National Institute of General Medical Sciences award number R35GM128648 (for the in-lab developments), the National Center for Advancing Translational Sciences award number 5TL1TR002318-08 (for support of LGB), and through the National Institute of Environmental Health Sciences to the University of Washington EDGE Center award number P30ES007033. The REDCap used for human subjects consent, enrollment, and survey is supported by the University of Washington Institute of Translational Health Sciences, which is funded by the National Center for Advancing Translational Sciences under award number UL1TR002319. The content is solely the responsibility of the authors and does not necessarily represent the official views of the National Institutes of Health or other funding bodies.

