## Supplementary Information for "A Flexible and Responsive Remote Study Design to Assess Gene Expression Changes During Wildfire Smoke Exposure with homeRNA, an At-home Blood Sampling Kit"

### Table of Contents:

| | |
|---|---|
| **Supplemental Figures** | S2 – S4 |
| **Supplemental Table** | S5 |
| **Appendix 1: homeRNA kit components and instructions** | S6 – S7 |
| **References** | S7 |
| **Instructions for Use (IFU)** | S8 – S9 |
| **Screening REDCap Survey** | S10 |
| **Eligibility REDCap Survey** | S11 |
| **Enrollment REDCap Survey** | S12 – S17 |
| **Baseline REDCap Survey** | S18 – S26 |
| **Sample Collection REDCap Survey** | S27 – S31 |
| **Smoke Exposure REDCap Survey** | S32 – S38 |
| **Closing REDCap Survey** | S39 – S42 |
| **Follow-up REDCap Survey** | S43 – S45 |

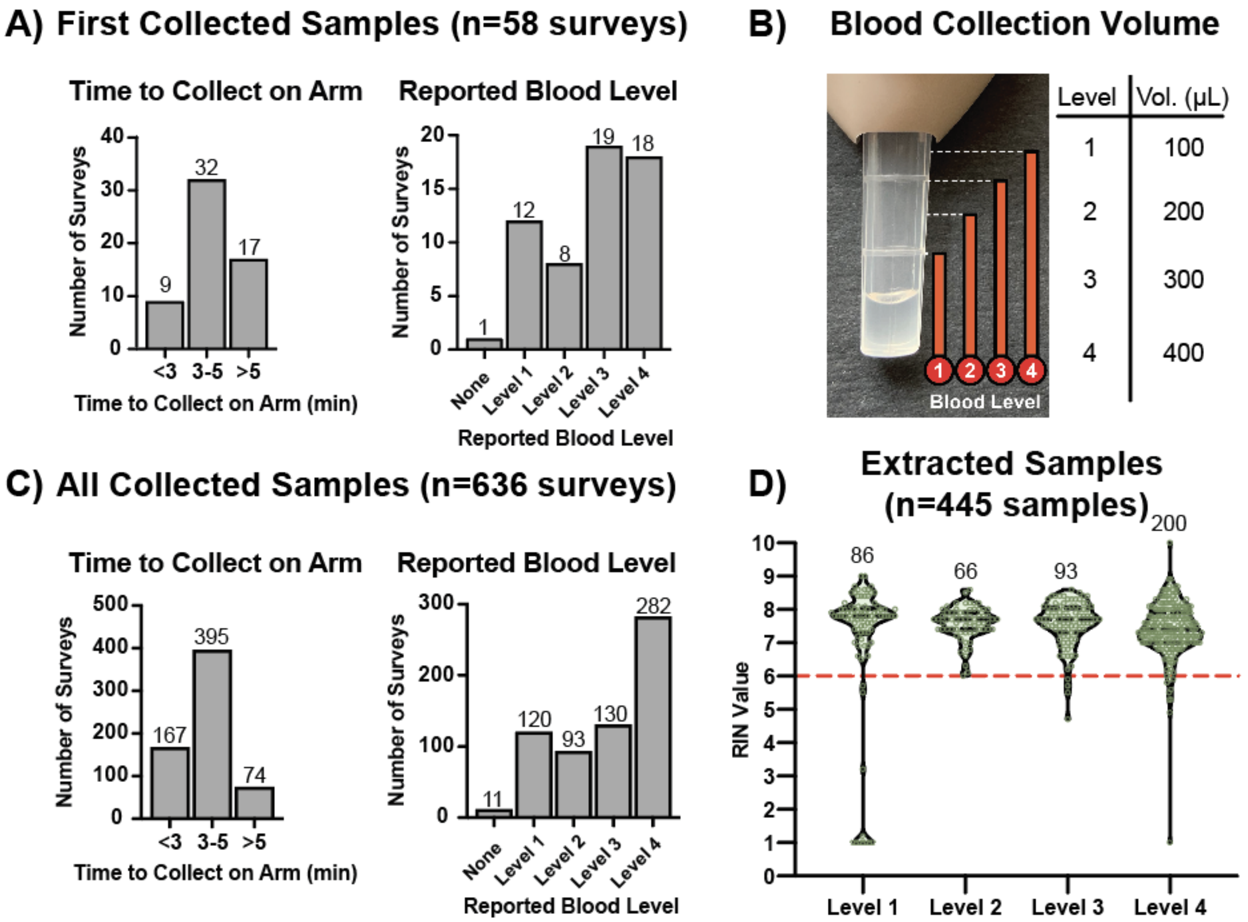


**Figure S1.** Blood collection volume associated with the Tasso-SST device had little effect on resulting RNA Integrity (RIN) values. Participant survey responses on the total time to collect blood on the upper arm with the Tasso-SST device (left) and the reported blood level collected (right) from A) first collected sample (n=58 surveys) and C) all collected samples (n=636 surveys) from 58 participants. B) Image of Tasso-SST blood collection device with approximate volume levels marked. Participants were asked to report blood level by eye after blood collection based on this image. D) Distribution of RIN values from reported blood collection levels (n=455 samples from the 37 participants that had samples extracted). Seven samples were reported to have no blood collected, so those were excluded here. RIN values are given on a scale from 1 to 10, with a RIN of 1 representing the most degraded RNA and a RIN of 10 representing the most intact RNA.

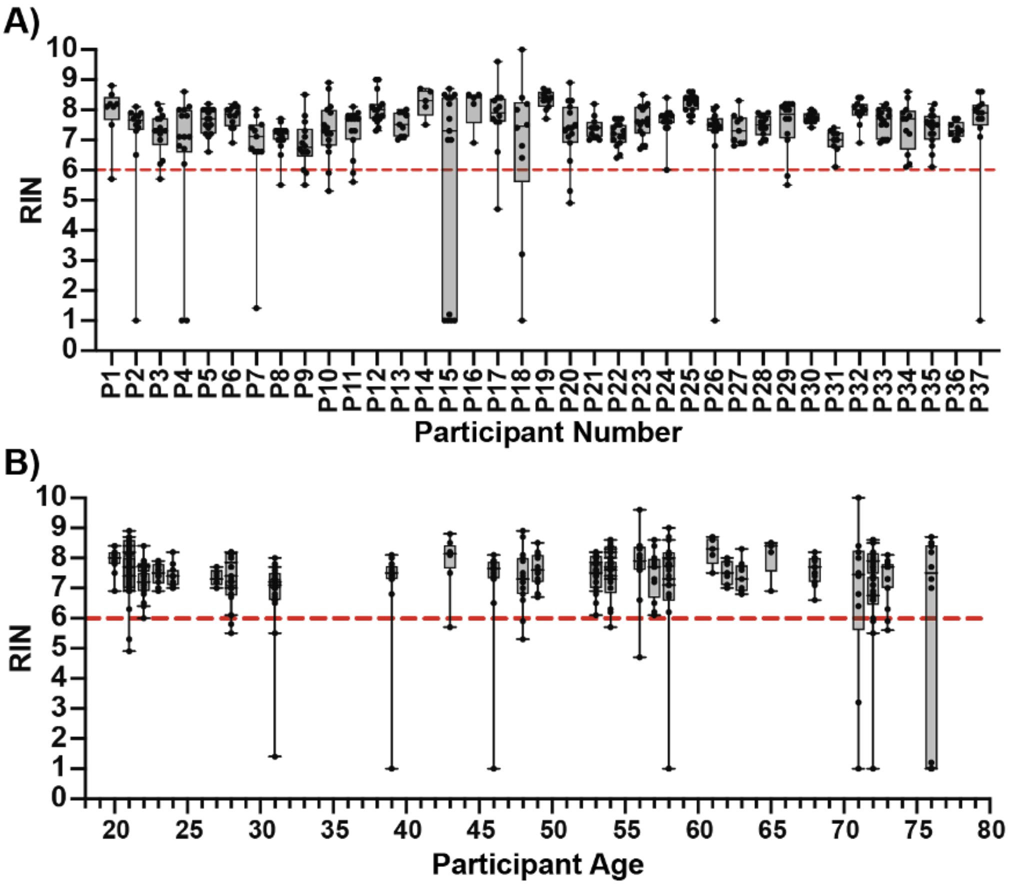


**Figure S2**. RNA stability of extracted homeRNA-stabilized samples based on each individual participant and participant age. A) Distribution of RNA Integrity Number (RIN) values from each participant (n=37 participants) from which samples were extracted. B) Distribution of RIN values according to participant age (n=37 participants). RIN values are given on a scale from 1 to 10, with a RIN of 1 representing the most degraded RNA and a RIN of 10 representing the most intact RNA.

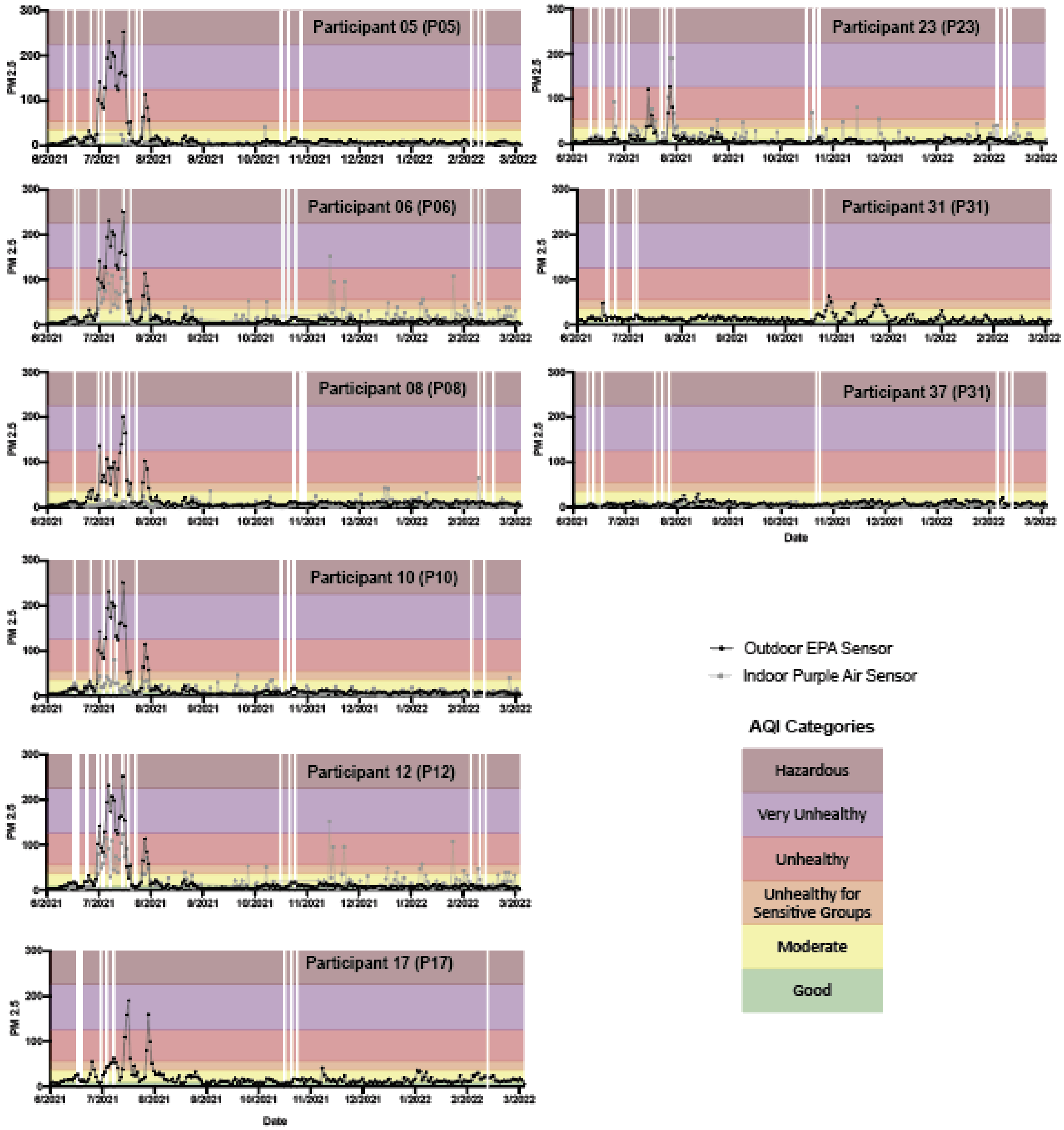


**Figure S3.** PM2.5 data for participants that were analyzed with the Nanostring Autoimmune gene panel. Data from participants exposed to high (left), medium (right, top), or low (right, bottom two) levels of wildfire smoke, plotted against the homeRNA sampling timepoints (white vertical lines with Tasso symbols on top), outdoor EPA sensor (black lines with data points), indoor PurpleAir sensor (grey lines with data points), and AQI categories as defined in Table S1.

**Table S1**. Air Quality Index (AQI) Categories and $PM_{2.5}$ Cutoffs as Defined by the Environmental Protection Agency updated on February 2024 (airnow.gov/aqi)

| AQI Category | $PM_{2.5}$ ($ug/m^3$) Breakpoints for each AQI Category |
|---|---|
| Good | 0.0 - 9.0 |
| Moderate | 9.1 - 35.4 |
| Unhealthy for Sensitive Groups | 35.5 - 55.4 |
| Unhealthy | 55.5 - 125.4 |
| Very Unhealthy | 125.5 - 225.4 |
| Hazardous | ≥225.5 |

**Appendix 1:** *home*RNA kit components and instructions (all materials reprinted from Haack, Lim *et al.* SI with permission from Haack, Lim *et al. home*RNA: A Self-Sampling Kit for the Collection of Peripheral Blood and Stabilization of RNA. *Anal. Chem.* 2021, 93, 39, 13196–13203. Copyright 2021 American Chemical Society. for ease of reference) (1).

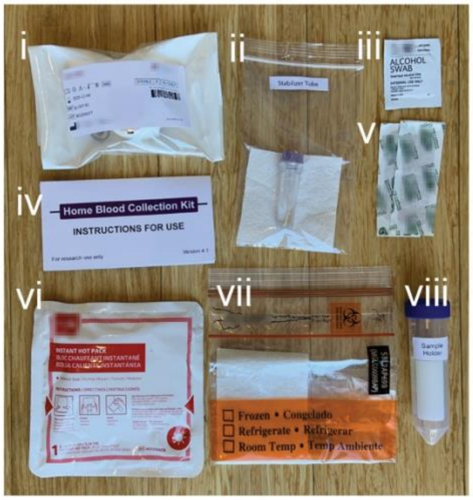


**Figure A1.** Components of the *home*RNA kit. Kit components include i) the Tasso-SST™ device, ii) the stabilizer tube containing RNA*later*™ iii) alcohol wipes iv) instructions for use, v) sterile bandages, vi) hot pack for warming the arm prior to application of the Tasso-SST™, vii) sample return bag and viii) sample holder with 3D printed insert to hold the sample tube in place.

**Table A1.** Components of the *home*RNA blood kit.

| Kit Component | Manufacturer(s) | Quantity |
|---|---|---|
| Sterile Tasso-SST™ blood collection device | Tasso, Inc. | 1 |
| RNA stabilizer tube | Our Lab | 1 |
| Instant heat pack | Medline Industries, Inc. | 1 |
| Sterile Alcohol Wipe | Covidien, BD | 2 |
| Sterile bandage | Band-Aid, Curad | 1 |
| Specimen transport bag with absorbent pad | Minigrip | 1 |
| 50 mL conical tube with stabilizer tube insert | BD, Our Lab | 1 |
| Instructions for use | Our Lab | 1 |
| Blood-stabilizer mixing instruction card | Our Lab | 1 |

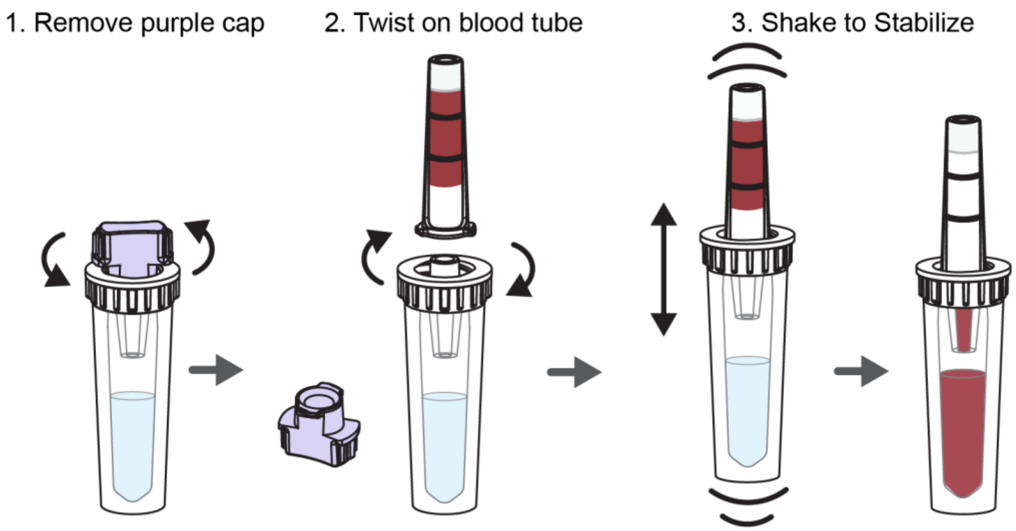


**Figure A2.** Schematic workflow of the stabilization process. The steps for stabilizing a blood sample collected with the Tasso-SST™ are as follows: 1) removal of the purple cap, 2) twisting on the Tasso-SST™ blood tube with the blood sample, 3) shaking up and down vigorously to mix the sample with the stabilizer resulting in RNA stabilized blood.

**Instructional video for *home*RNA**

An instructional video for using *home*RNA can be found at the following link:
https://youtu.be/iV3GZ8SmmuM

#### PREPARE DEVICES AND APPLICATION SITE

**FIRST TIME USERS**

Please watch the instructional video provided at

https://www.you-tube.com/watch?v=iV3GZ8SmmuM

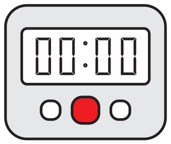


**1. Wash hands and get a timer.**

You will use a timer in Steps 2 and 10.

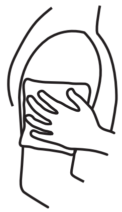

**2. Apply hot pack to upper arm for 2 minutes.**

Warming helps your blood flow better.

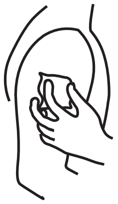

**3. Clean arm with alcohol wipe.**

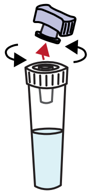

**4. Open the stabilizer tube by twisting off the purple cap.**

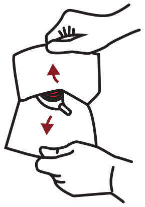

**5. Open Tasso pouch by pulling apart white and clear layers.**

Discard cap in pouch.

#### COLLECT BLOOD USING TASSO-SST

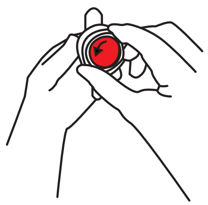

**6. Remove clear plastic cover over the red button.**

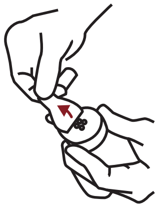

**7. Peel paper tab behind the red button.**

Keep the tube pointing down.

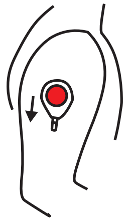

**8. Stick device to shoulder.**

Do not remove once it is on.

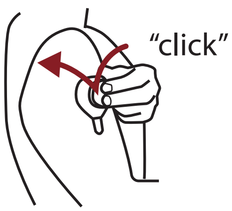


**9. Press button quickly and firmly until it can't go any farther. Wait 2 seconds then let go.**

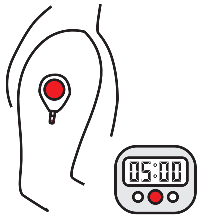


**10. Start a 5 minute timer.**

Keep arm at your side. You wont see blood right away. It can take up to a minute for blood to flow.

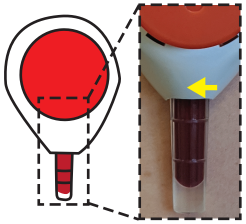

**11. After 5 minutes or when the tube fills, *whichever comes first*, peel off the device.**

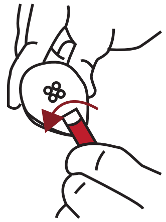

**12. Remove tube by firmly twisting a quarter turn and pulling down.**

This may take a bit of finger strength.

#### MIX AND PACKAGE

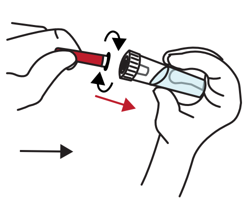

**13. Bring together the blood tube and stabilizer tube and screw these together tightly.**

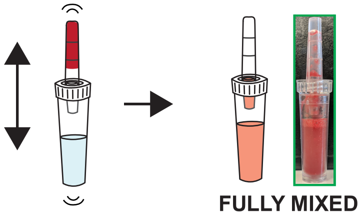


**14. With the stabilizer tube on the bottom, shake hard up and down to mix. Stop when mixed.**

Some fluid may remain in blood tube. When mixed, the color is the same.

**DO NOT DISCONNECT TUBES.**

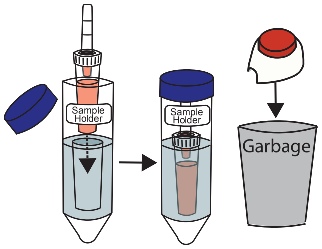


**15. Place sample in sample holder. Throw away used Tasso device.**

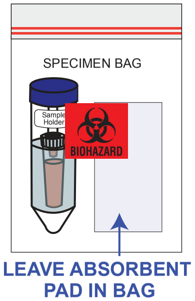


**16. Place blood sample in the specimen bag. Wash your hands with soap and water**

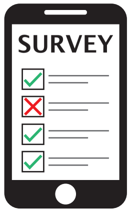


**17. Fill out the online Collection Survey that was emailed to you.**

You will need the Unique Code on the box.

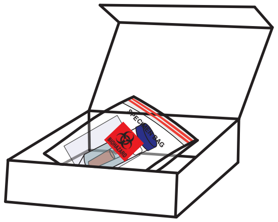

**18. After filling out the survey, place your sample back in the box, and return using the provided shipping return bag.**

For research use only

### INSTRUCTIONS FOR USE

#### Home Blood Collection Kit

If you need assistance please contact us at _study email_ or call _study phone_

**Thank you for participating in our study!**

#### KIT CONTENTS

Make sure your kit contains all components listed below

**1) Tasso-SST device**

**2) Stabilizer tube**

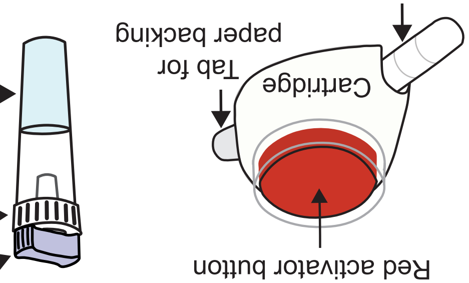


**3) Alcohol wipes**

**4) Hot pack**

**5) Bandage**

**6) Specimen bag**

**7) Sample Holder**

**8) Mail return bag**

#### INTENDED USE

TASSO-SST is a single use blood collection device that is intended for the self collection of capillary blood from the upper arm of adults (18 years or older). The stabilizer tube contains liquid that is intended for stabilizing the collected blood. The home blood sampling kit is for academic research use.

#### STORAGE

Store at 15 - 30ºC (60 - 80ºF) in a dry place.

#### WARNINGS

- The TASSO-SST is a sterile device. Do not open until use.
- The TASSO-SST device contains sharps. Handle with care.
- For external use only.
- Keep out of reach of children.
- Use while seated as fainting may occur during blood sampling procedure.
- Do not activate the red button until device is firmly on skin.
- Wipe application site with alcohol wipe to reduce infection risk.
- Always use a new unopened pouch of Tasso-SST. Do not re-use.

#### ONLINE SYMPTOM AND COLLECTION SURVEY

Please check your e-mail or text message based on the preference you has listed for a link to fill out the daily use online survey.
If you cannot find the link, e-mail us at _study email_

### Immune Response to Wildfire Exposure Study: Screening Survey

Thank you for your interest in our study!

Please complete the survey below to determine if you are eligible for participation.

To learn more information about the study, please visit our website at http://depts.washington.edu/bcmelab/wildfires/

Thank you!

#### Please fill out the information below.

| | |
|---|---|
| Are you currently 18 years old or older? | ◯ Yes<br>◯ No |
| Do you live in an area that is prone to wildfires or wildfire smoke exposure?<br><br>Eligible areas include:<br><br>Alaska, Arizona, California, Colorado, Florida, Idaho, Montana, Nevada, New Mexico, Oklahoma, Oregon, Texas, Utah, Washington, and Wyoming. | ◯ Yes<br>◯ No |
| Do you live in an area that typically experiences year-round or prolonged (several months) exposure to smoke, either from wildfires, controlled burns or other wood-burning activities (wood-burning stoves or campfires)? | ◯ Yes<br>◯ No |
| Are you pregnant? | ◯ Yes<br>◯ No |
| Are you currently residing in a correctional facility? | ◯ Yes<br>◯ No |
| Are you a friend or family member of the researchers conducting this study? | ◯ Yes<br>◯ No |
| What is your current residential zipcode? | ______________________ |
| Area | ______________________ |
| What is your email address? Please note the email address you provide here will be used to contact you throughout the study. | ______________________ |
| Are you part of the Clean Air Ambassador Program from Clean Air Methow? | ◯ Yes<br>◯ No |

| | |
|---|---|
| What is your name? | ____________________________________ |
| Does your occupation require you to work outside? | ◯ Yes<br>◯ No<br>◯ Sometimes |
| What is your occupation? | ____________________________________ |
| Do you have a PurpleAir Indoor air monitor installed in your living space?<br><br>Please note this is not required for enrollment. | ◯ Yes<br>◯ No |
| Is your home connected to WiFi? | ◯ Yes<br>◯ No |
| We will be providing PurpleAir Indoor air monitors to a subset of study participants. If you are selected for this sub-group would you be willing to install a WiFi equipped PurpleAir indoor air quality monitor in your living space and sharing the data collected from this air monitor with the study team?<br><br>Please note, you can still participate in the study without agreeing to install a PurpleAir indoor air quality monitor.? | ◯ Yes<br>◯ No |
| In this study, we will be asking you to collect between 3 and 15 blood samples during and after wildfire season. The study team will direct you when to collect blood samples and how many to collect. The timing of these samples cannot be predicted as they will depend on the occurrence of wildfire events. Would you feel comfortable participating in this study given the unpredictable nature of the sampling timeline? | ◯ Yes<br>◯ No |

We have provided a link below to the consent form for this study to give more information about the study procedures and what to expect from participation in this study. You do not need to sign this form now. If selected to participate in the study, you will be emailed a link to the consent form to sign online.

[Attachment: "Consent Form.pdf"]

Press submit to be added to the waiting list for this study.

### Enrollment Survey

Please complete the survey below.

Thank you!

| | |
|---|---|
| First Name | __________________ |
| Last Name | __________________ |
| Preferred Name | __________________ |
| Assigned sex at birth | ◯ Male<br>◯ Female |
| Phone number | __________________<br>(Include Area Code) |
| Can this phone number receive texts? | ◯ Yes<br>◯ No |
| Address Line 1 | __________________ |
| Address Line 2 (Optional) | __________________ |
| City | __________________ |
| State | __________________ |
| ZIP Code | __________________ |
| Your completed homeRNA kits will be picked up by UPS at a scheduled pick up time.?<br><br>Please indicate where you would like your?completed kit picked up | ◯ Front door/Porch<br>◯ Back door/Porch<br>◯ Garage<br>◯ Reception/Lobby<br>◯ Office<br>◯ Mail room |

**The following health related questions will help us understand your specific immune response to Wildfire smoke exposure.**

What is your height in inches?

__________________________________
(For your reference, 60 inches is 5 feet)

What is your weight in pounds?

__________________________________

Has your doctor or medical provider ever told you you have any of the following diseases? (Check all that apply)

- ☐ Cystic Fibrosis
- ☐ Asthma
- ☐ Lung Disease
- ☐ Hypertension/High Blood Pressure
- ☐ Hyperlipidemia/Hypercholesterolemia/High Cholesterol
- ☐ Other Heart Disease
- ☐ Anemia
- ☐ Liver Disease
- ☐ Diabetes
- ☐ Cancer
- ☐ Chronic Kidney Disease
- ☐ Other Chronic Condition
- ☐ None of the above

Please select all lung diseases that apply to you.

- ☐ Chronic Obstructive Pulmonary Disorder (COPD)
- ☐ Pulmonary Fibrosis or Idiopathic Pulmonary Fibrosis
- ☐ Bronchiectasis
- ☐ Alpha-1Antritrypsin Deficiency
- ☐ Other Lung Disorder

If you selected "other" above, please describe your lung disease.

__________________________________

Please select all heart diseases that apply to you.

- ☐ Congenital Heart Disease
- ☐ Coronary Artery Disease or History of Heart Attack
- ☐ Congestive Heart Failure
- ☐ Other

If you selected "other" above, please describe your heart disease.

__________________________________

If you selected "other chronic condition" above, please specify here (optional).

__________________________________

Have you ever had an organ transplant?

- ◯ Yes
- ◯ No

If yes, which organ?

__________________________________

| | |
|---|---|
| Have you ever been diagnosed with an autoimmune condition? If so, please check all that apply. | ☐ I have not been diagnosed with an autoimmune condition.<br>☐ Autoimmune Thyroid Disease (i.e., Hashimoto's thyroiditis)<br>☐ Lupus<br>☐ Multiple Sclerosis<br>☐ Cytopenia<br>☐ Colitis/Inflammatory Bowel Disease<br>☐ Other |
| If you selected "other", please describe your autoimmune condition. | __________________________________ |
| Have you ever been diagnosed with any other immune related condition? (Check all that apply) | ☐ I have not been diagnosed with any other immune related conditions<br>☐ Periodic/Frequent Fevers<br>☐ Immune Deficiency<br>☐ Eczema |
| Please specify your immune deficiency disease (Optional). | __________________________________ |
| Do you have seasonal allergies or hay fever? | ◯ Yes<br>◯ No |
| Is there a time of the year in which your allergies are worse? | ◯ Yes<br>◯ No |
| If yes, please specify the season. | ☐ Spring<br>☐ Fall<br>☐ Winter<br>☐ Summer<br>☐ I Don't Know |
| What are your typical seasonal allergy symptoms? | ☐ Runny nose<br>☐ Stuffy nose<br>☐ Itchy mouth or throat<br>☐ Sneezing<br>☐ Wheezing<br>☐ Shortness of breath<br>☐ Cough<br>☐ Rashes<br>☐ Fatigue<br>☐ Headache<br>☐ Nausea or Vomiting<br>☐ Fever<br>☐ Other<br>☐ None of the above |
| If other, please specify your symptoms. | __________________________________ |

S14

| | |
|---|---|
| Do you currently take any non-steroidal anti-inflammatory agents (NSAIDS) at least once a week, with or without a perscription? These include<br><br>- ibuprofen (Motrin, Advil)<br>- naproxen (Naprosyn, Aleve, Anaprox, Naprelan)<br>- diclofenac (Cambia, Cataflam, Voltaren, Zipsor)<br>- indomethacin (Indocin)<br>- diflunisal (Dolobid)<br>- etodolac (Lodine, Lodine XL)<br>- ketoprofen (Orudis, Orudis KT, Oruvail)<br>- ketorolac (Acular, Acular LS, Acular PF, Acuvail)<br>- nambumetone (Relafen)<br>- oxaprozin (Daypro)<br>- piroxicam (Feldene)<br>- salsalate (Disalate)<br>- sulindac (Clinoril)<br>- tolmetin (Tolectin 600, Tolectin DS)<br>- celecoxib (Cerebrex)) | ◯ Yes<br>◯ No |
| Do you take aspirin at least once a week, with or without a perscription? | ◯ Yes<br>◯ No |
| Do you currently take any other medications (including over the counter medications) at least once a week? | ◯ Yes<br>◯ No |
| Are you taking any of the following medications? Select all that apply. | ☐ ACE Inhibitors (e.g. benazepril (Lotensin), captopril (Capoten), enalapril (Epaned, Vasotec), fosinopril (Monopril), lisinopril (Prinivil, Zestril))<br>☐ Angiotensin Receptor Blockers (ARBs) (e.g. losartan (Cozaar), valsartan (Diovan), irbesartan (Avapro), candesartan (Atacand), telmisartan (Micardis), Olmesartan (Benicar), others)<br>☐ Beta-Blockers (e.g. metoprolol (Lopressor, Toprol XL), atenolol (Tenormin), carvedilol (Coreg, Coreg CR), others)<br>☐ Other blood pressure medications<br>☐ I am not taking any of these medications |
| If other, please specify your blood pressure medication. | __________________________ |
| Are you taking any of the following cholesterol medications? | ☐ Statins (e.g. atorvastatin (Lipitor), rosuvastatin (Crestor), simvastatin (Zocor), pravastatin (Pravachol), lovastatin (Altoprev, Mevacor), fluvastatin (Lescol, Lescol XL), pitavastatin (Livalo))<br>☐ Other (e.g. ezetimibe (Zetia), fenofibrate (Antara, Fenoglide, Lipofen, others), others)<br>☐ I am not taking any of these medications |
| If other, please specify your cholesterol medication. | __________________________ |

| | |
|---|---|
| Do you take any other medications at least once a week? If yes, please select all that apply. | ☐ Oral Corticosteroids (e.g. Prednisone (Deltasone, Prednicot, others))<br>☐ Inhaled Corticosteroids (e.g. fluticasone (Flovent), beclomethasone (QVar))<br>☐ Inhaled Bronchodilators (e.g. albuterol (Accuneb, ProAir HFA, Proventil, others))<br>☐ Other Asthma Medications<br>☐ Diabetes Medications<br>☐ Anti-TNF Medications (e.g. infliximab (REMICADE), adalimumab (Humira), certolizumab (Cimzia), golimumab (Simponi), etanercept (Enbrel, Erelzi), others)<br>☐ IL-6 Pathway Inhibitors (e.g. sarilumab (Kevzara), tocilizumab (Actemra), siltuximab (Sylvant), others)<br>☐ Conventional Disease-Modifying Anti-Rheumatic Drugs (DMARDs) (e.g., cyclosporine (Neoral, Sandimmune, Gengraf), cyclophosphamide (Cytoxan, Neosar), hydroxychloroquine (Plaquenil), leflunomide (Arava), methotrexate (Trexall, Otrexup, Rasuvo), mycophenolate (CellCept, Myfortic, others), sulfasalazine (Azulfidine))<br>☐ JAK Inhibitors (baricitinib (Olumiant), ruxolitinib (Jakafi), fedratinib (Inrebic), tofacitinib (Xeljanz))<br>☐ Blood Thinning Medications (e.g. warfarin (Coumadin), heparin, enoxaparin (Lovenox), apixaban (Eliquis), rivaroxaban (Xarelto))<br>☐ Platelet Inhibitors (e.g. clopidogrel (Plavix), prasugrel (Effient), ticagrelor (Brilinta))<br>☐ Thyroid Medications (e.g. levothryroxine (Levothroid, Levoxyl, Synthroid))<br>☐ Other (Prescribed/Non-Prescribed/Vitamins or Supplements)<br>☐ I am not taking any of these medication |
| If you selected "other asthma medications" above, please specify your asthma medication. | ______________________ |
| Please specify your diabetes medication. | ______________________ |
| Please specify any additional medications you take at least once a week, both prescribed or not prescribed. | ______________________ |
| Have you ever been diagnosed with COVID-19? | ○ Yes<br>○ No |
| If yes, please specify the approximate date(s) of COVID-19 illness. | ______________________ |
| Do you have any long COVID-19 symptoms? | ○ Yes<br>○ No |
| If yes, please specify your long COVID-19 symptoms. | ____________________________ |
| Did you receive a COVID-19 vaccination? | ○ Yes<br>○ No<br>○ I do not remember |

| | |
|---|---|
| If yes, please specify which COVID-19 vaccine you received. | ______________________ |
| If yes, please specify the approximate date of when you received the first dose. | ______________________ |
| If yes, please specify the approximate date of when you received the second dose (if applicable). | ______________________ |
| Did you receive a flu vaccine within the last 6 months? | ◯ Yes<br>◯ No<br>◯ I do not remember |
| If yes, please specify the approximate date of the flu vaccination. | ______________________ |
| Have you received any other immunizations within the last 6 months? | ◯ Yes<br>◯ No |
| If yes, please specify which immunization you received. | ______________________ |
| If yes, please specify the approximate date when you received your last vaccination. | ______________________ |
| Do you have any vaccinations scheduled or do you plan to schedule any within the next 6 months? | ◯ Yes<br>◯ No |
| If yes, please specify which immunization you have scheduled or plan to schedule. | ______________________ |
| Please specify the approximate date you plan to receive this immunization. | ______________________ |

### Baseline Survey

Please complete the survey below.

Thank you!

---

Please enter the sample code of the kit you have just finished using: ______________________

The code will be located on the outside of the individual kit box and will have the format of XXXX - X?

---

Please indicate the date and time you collected your sample.

---

Date: ______________________

---

Time: ______________________

---

You should have been provided a temperature and humidity monitor in the first package we sent you located in the extra materials box. Using this monitor, please report the temperature and humidity below.

---

Temperature (Farenheit): ______________________

---

Humidity (%): ______________________

---

Did you have any challenges setting up the PurpleAir indoor air monitor?
- ◯ Yes
- ◯ No

---

Please describe the issues you had setting up your PurpleAir indoor air monitor (optional): ______________________

---

What is the name you have given your PurpleAir sensor? ______________________

---

Have you registered your PurpleAir monitor as public or private?
- ◯ public
- ◯ private

---

Please copy and paste the unique link associated with your PurpleAir sensor's data into the field below. This link was sent by PurpleAir to the email address you provided during your PurpleAir device registration. ______________________

---

What is the PM 2.5 reading of the nearest outdoor PurpleAir monitor? ______________________

---

What is the current PM 2.5 reading on the PurpleAir indoor air monitor? ______________________

**We will now ask a few questions about your health:**

What is your date of birth? __________________________________

Have your medications changed since you last filled out the enrollment survey?
- ◯ Yes
- ◯ No

Please select from the following:
- ◯ I started taking a new medication
- ◯ I stopped taking a medication
- ◯ My medications have not changed since I last filled out the survey

Please describe the new medication: __________________________________

Please describe which medication you stopped taking: __________________________________

Have you had a vaccination in the past 2 weeks?
- ◯ Yes
- ◯ No

Please select which vaccination you received in the last two weeks.
- ☐ Flu
- ☐ COVID-19 (Pfizer, Moderna or Johnson and Johnson)
- ☐ Other

If you selected "other" please describe the vaccination you have received. __________________________________

Do you plan to have any vaccinations in the next month?
- ◯ Yes
- ◯ No
- ◯ I don't know

If yes, which vaccination are you planning to receive? __________________________________

Approximately when do you plan to have this vaccine? __________________________________

Has your address changed since you last filled out a survey?
- ◯ Yes
- ◯ No

If yes, what is the zipcode of your new address? __________________________________

Do you anticipate your address will change in the next 2 months?
- ◯ Yes
- ◯ No

#### We will now ask some questions about your experience using the homeRNA blood kit today:

Approximately how long (in minutes) did it take to use the homeRNA blood kit today?

◯ Less than 5 minutes
◯ 5-10 minutes
◯ 11-15 minutes
◯ 16-20 minutes
◯ More than 20 minutes

While using the homeRNA blood kit today, approximately how long (in minutes) did you leave the Tasso-SST blood collection device on your arm?

◯ Less than 3 minutes
◯ 3 - 5 minutes
◯ More than 5 minutes
◯ I am unsure

Based on the image below, approximately how much blood did you collect today? Choose the closest level.

◯ I was unable to collect any blood
◯ Level 1
◯ Level 2
◯ Level 3
◯ Level 4

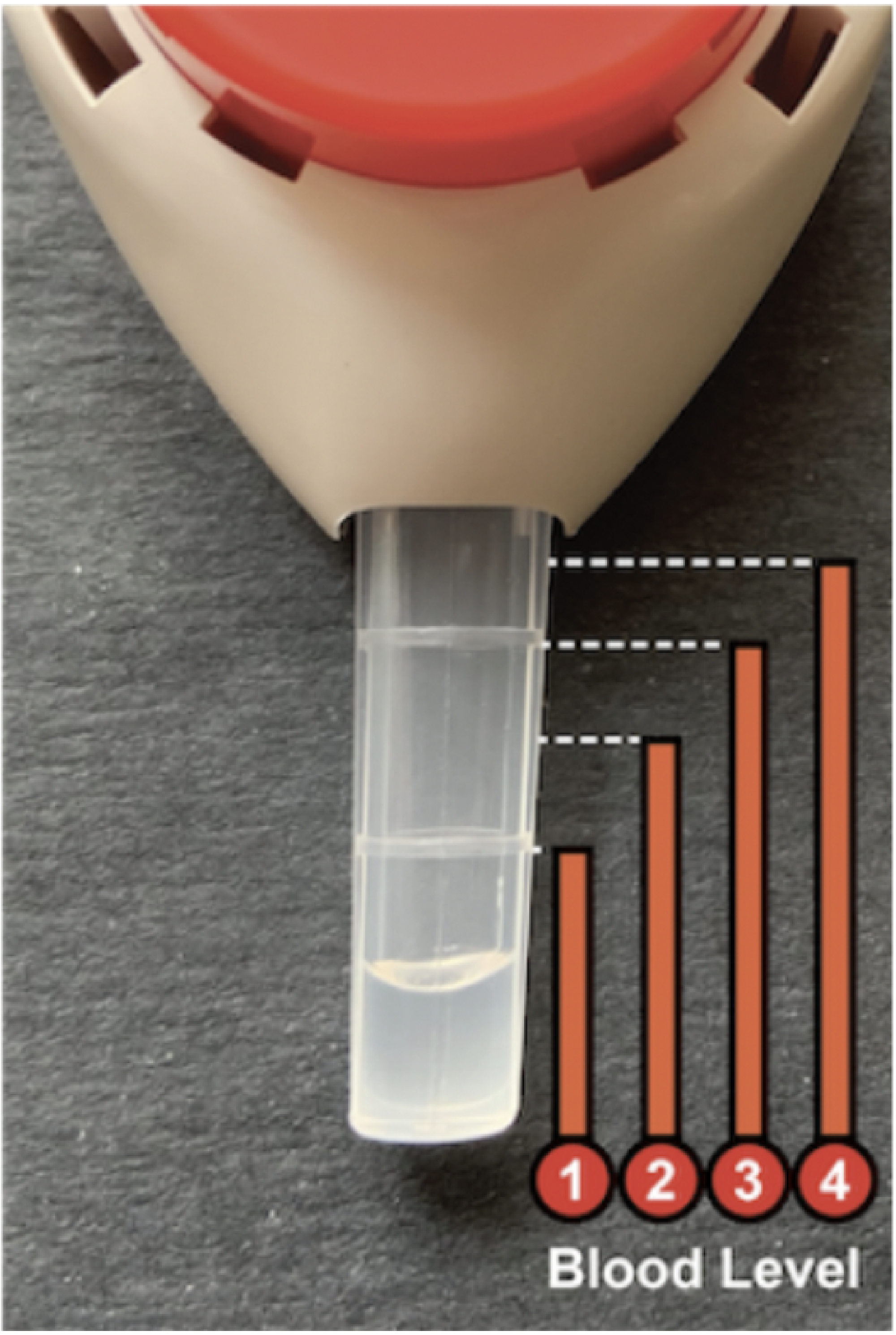


| | |
|---|---|
| Did you experience any pain while using the home RNA blood kit today? | ◯ No pain<br>◯ Mild pain<br>◯ Moderate pain<br>◯ Severe pain<br>◯ Very severe pain |
| Has the temporary pain resolved? | ◯ Yes<br>◯ No |

#### How easy was it to use the components of the kit listed below?

| | Very difficult | Somewhat difficult | Neither difficult nor easy | Somewhat easy | Very easy |
|---|---|---|---|---|---|
| The Tasso-SST device | ◯ | ◯ | ◯ | ◯ | ◯ |
| The blood stabilization tube | ◯ | ◯ | ◯ | ◯ | ◯ |

Did you experience any issues with the Tasso-SST blood collection device?

◯ Yes
◯ No

Please describe any issues you had with the Tasso-SST blood collection device. (Optional)

__________________________________

Did you experience any issues with the stabilizer tube or mixing your samples?

◯ Yes
◯ No

Please describe any issues you had with the stabilizer tube or mixing your samples. (Optional)

__________________________________

Please use the space for any other comments on your experience today collecting blood samples using the home RNA blood kit. (Optional)

__________________________________

Please briefly describe where you plan to leave your completed kit before it is picked up??

e.g. I will leave it inside until tomorrow morning where I will place it on the porch for pickup.

__________________________________

#### We will now ask you some questions related to wildfire smoke exposure and related stress.

Have you had prolonged exposure (>30 min) to smoke (i.e. from a wildfire, campfire, controlled burn in your area) in the past 6 months?

◯ Yes
◯ No

If yes, please briefly describe:

__________________________________

In the past year, how many weeks were you exposed to poor air quality (e.g. wildfire smoke, controlled burns, wood burning stove, other pollution)?

◯ I was not exposed to any poor air quality in the past year
◯ I was exposed 1 day to 2 weeks of poor air quality
◯ I was exposed to 2 - 4 weeks of poor air quality
◯ I was exposed to greater than 4 weeks of poor air quality
◯ I am not sure how long I was exposed to poor air quality

Which of the following contributed to low air quality that you were exposed to in the past year?

☐ Wildfire
☐ Controlled burns
☐ Campfires
☐ Wood burning stove
☐ Industry
☐ Agriculture
☐ Other pollution

Do you currently have any of these physical symptoms?

- ☐ Coughing
- ☐ Trouble breathing normally
- ☐ Stinging eyes
- ☐ A scratchy throat
- ☐ Runny nose
- ☐ Irritated sinuses
- ☐ Wheezing and shortness of breath
- ☐ Chest pain
- ☐ Headaches
- ☐ Asthma attack
- ☐ Tiredness
- ☐ Fast heartbeat
- ☐ Other
- ☐ I do not have any of these physical symptoms

If other, please describe your symptoms: __________________________________

Are you currently feeling sick (e.g., cold or flu)?

- ◯ Yes
- ◯ No

Please briefly describe your symptoms related to feeling sick. __________________________________

| | I was experiencing less stress than normal | | I was experiencing about the same amount of stress as normal | | I was experiencing more stress than normal |
|---|---|---|---|---|---|
| Compared to your normal level of stress, how stressed were you today? | ◯ | ◯ | ◯ | ◯ | ◯ |

| | I generally have the same amount of stress as normal | | I am somewhat more stressed compared to normal | | I can become extremely stressed compared to normal |
|---|---|---|---|---|---|
| Compared to your normal level of stress, how would you say you react to decreased air quality in your physical environment? | ◯ | ◯ | ◯ | ◯ | ◯ |

| | I would feel very comfortable using this kit during a natural disaster | | I would feel neutral about using this kit during a natural disaster | | I would feel very uncomfortable using this kit during a natural disaster |
|---|---|---|---|---|---|

How comfortable do you think you would be using this kit in the midst of a natural disaster, assuming you are not in any immediate danger? (e.g. wildfire, earthquake, hazardous weather) ○ ○ ○ ○ ○

**We will now ask you a few questions related to your daily activities and stress levels.**

What is your occupation? ______________________

About how much time (in hours) do you spend outside per day? ______________________

About how much time (in hours) do you spend in a bus or car per day? ______________________

Are you regularly exposed to environmental stressors such as poor air quality, extreme temperatures, physical injuries, exposures to chemicals, etc.?
○ Yes ○ No

If yes, please specify: ______________________

Approximately how many hours of sleep do you get each night on average?
○ Less than 6 hours
○ 6 - 9 hours
○ 9 - 12 hours
○ More than 12 hours

What activities do you engage in on a regular basis? Check all that apply.
☐ Walking
☐ Running
☐ Swimming
☐ Yoga
☐ Weights
☐ Dancing
☐ Biking
☐ Hiking
☐ Other
☐ I do not engage in exercise on a regular basis

If other, please specify: ______________________

On a typical week, how much time do you spend exercising? This includes both mild and moderate exercise, such as walking, as well as intense exercise such as running.
○ 0 to 30 minutes
○ 30 to 60 minutes
○ 1 to 2 hours
○ 2 to 5 hours
○ >5 hours

Do you regularly participate in one or more of these mindfulness activities? Check all that apply.

- ☐ Meditation
- ☐ Deep breathing
- ☐ Body scan
- ☐ Visualization
- ☐ Prayer
- ☐ Religious/faith service
- ☐ Other
- ☐ I do not participate in mindfulness activities on a regular basis

If other, please specify: ______________________

How many hours per week would you say you spend on these mindfulness activities? ______________________

| | They do not help reduce my stress level | | They moderately help reduce my stress level | | They significantly help reduce my stress level |
|---|---|---|---|---|---|
| How well would you say these mindfulness activities reduce your stress levels? | ○ | ○ | ○ | ○ | ○ |

Are there any other hobbies that you enjoy regularly? (e.g. gardening, arts/crafts, video games, reading, writing, watching TV/movies, etc.)

- ○ Yes
- ○ No

If yes, please describe your hobbies. (Optional) ______________________

| | They do not help reduce my stress level | | They moderately help reduce my stress level | | They significantly help reduce my stress level |
|---|---|---|---|---|---|
| How well do these activities help to reduce your stress? | ○ | ○ | ○ | ○ | ○ |

| | It is not very important for managing my stress | | It is moderately important for managing my stress | | It is very important for managing my stress |
|---|---|---|---|---|---|
| How important is being able to go outside to managing your stress? | ○ | ○ | ○ | ○ | ○ |

| | I generally am not very stressed | | I have moderate levels of stress | | I am often very stressed |
|---|---|---|---|---|---|

Approximately how stressed would you say you are, on average? ○ ○ ○ ○ ○

Please use this space to write down anything else about your experience today. (Optional) ______________________

### Sample Collection Survey

Please complete the survey below.

Thank you!

---

Please enter the sample code of the kit you have just finished using: ____________________

The code will be located on the outside of the individual kit box and will have the format of XXXX - X?

---

Please indicate the date and time you collected your sample.

---

Date: ____________________

---

Time: ____________________

---

You should have been provided a temperature and humidity monitor in the first package we sent you. Using this monitor, please report the temperature and humidity below.

---

Temperature (Farenheit): ____________________

---

Humidity (%): ____________________

---

What is the current PM 2.5 reading of the PurpleAir Indoor air monitor? ____________________

---

What is the current PM 2.5 reading on the closest outdoor PurpleAir monitor? ____________________

---

Have your medications changed since you last filled out a survey?

- ◯ Yes
- ◯ No

---

Please select from the following:

- ◯ I started taking a new medication
- ◯ I stopped taking a medication
- ◯ My medications have not changed since I last filled out a survey

---

Please describe the new medication: ____________________

---

Please describe which medication you stopped taking: ____________________

---

Has your address changed since you last filled out a survey?

- ◯ Yes
- ◯ No

---

If yes, what is the zipcode of your new address? ____________________

Do you anticipate your address will change in the next 2 months?

◯ Yes
◯ No

**We will now ask you a few questions about how you are currently feeling:**

Do you currently have any of these physical symptoms?

☐ Coughing
☐ Trouble breathing normally
☐ Stinging eyes
☐ A scratchy throat
☐ Runny nose
☐ Irritated sinuses
☐ Wheezing and shortness of breath
☐ Chest pain
☐ Headaches
☐ Asthma attack
☐ Tiredness
☐ Fast heartbeat
☐ Other
☐ I do not have any of these physical symptoms

If you selected "other", please describe your symptoms:

__________________________________

Are you currently feeling sick (e.g., cold or flu)?

◯ Yes
◯ No

Please briefly describe your symptoms related to feeling sick.

__________________________________

| | I was experiencing less stress than normal | | I was experiencing about the same amount of stress as normal | | I was experiencing more stress than normal |
|---|---|---|---|---|---|
| Compared to your normal level of stress, how stressed were you today? | ◯ | ◯ | ◯ | ◯ | ◯ |

In the past week were you exposed to any environmental stressors such as poor air quality, extreme temperatures, physical injuries, exposures to chemicals, etc.?

◯ Yes
◯ No

If yes, please briefly describe:

__________________________________

Approximately how many hours of sleep per night did you get on average in the past week?

◯ Less than 6 hours
◯ 6 - 9 hours
◯ 9 - 12 hours
◯ More than 12 hours

Were you able to engage in your normal levels of exercise in the past week?

◯ Yes
◯ No
◯ I do not normally exercise

If not, please indicate any reasons you were unable to exercise, if applicable (optional):

__________________________________

Were you able to engage in your normal levels of mindfulness activities in the past week?

- ◯ Yes
- ◯ No
- ◯ I do not normally practice mindfulness

If not, please indicate any reasons you were unable to practice mindfulness, if applicable (optional):

__________________________________

Have you been able to engage in your other hobbies this past week? (e.g., gardening, arts/crafts, video games, reading, writing, watching movies or TV, etc.)

- ◯ Yes
- ◯ No
- ◯ I do not normally engage in other hobbies

If not, please indicate any reasons you were unable to engage in your other hobbies, if applicable (optional):

__________________________________

**We will now ask a few questions about your experience when using the homeRNA blood kit today:**

Approximately how long (in minutes) did it take to use the homeRNA blood kit today?

- ◯ Less than 5 minutes
- ◯ 5-10 minutes
- ◯ 11-15 minutes
- ◯ 16-20 minutes
- ◯ More than 20 minutes

While using the homeRNA blood kit today, approximately how long (in minutes) did you leave the Tasso-SST blood collection device on your arm?

- ◯ Less than 3 minutes
- ◯ 3 - 5 minutes
- ◯ More than 5 minutes
- ◯ I am unsure

Based on the image below, approximately how much blood did you collect today? Choose the closest level.

- ◯ I was unable to collect any blood
- ◯ Level 1
- ◯ Level 2
- ◯ Level 3
- ◯ Level 4

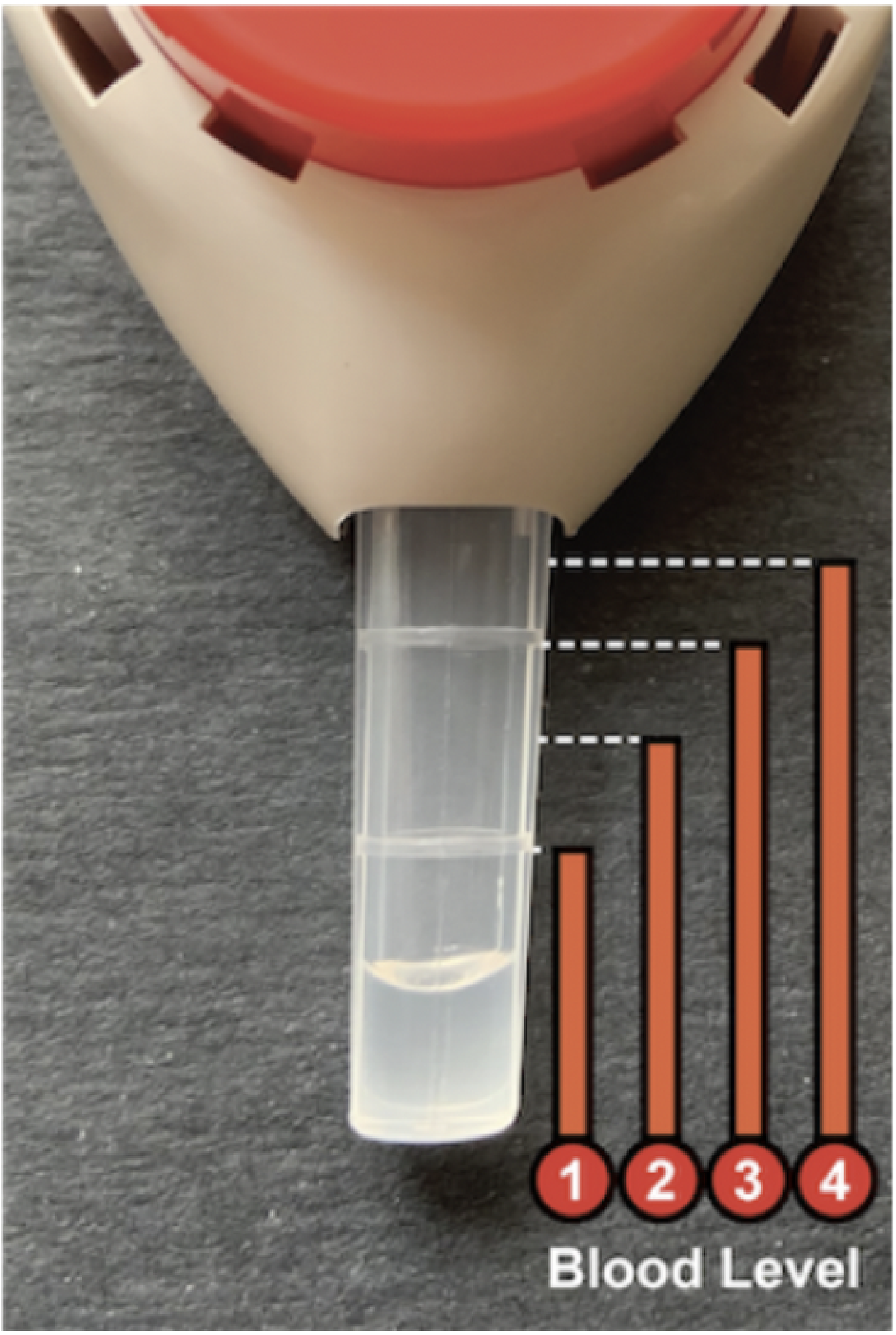


Did you experience any pain while using the home RNA blood kit today?

- ◯ No pain
- ◯ Mild pain
- ◯ Moderate pain
- ◯ Severe pain
- ◯ Very severe pain

Has the temporary pain resolved?

- ◯ Yes
- ◯ No

#### How easy was it to use the components of the kit listed below?

| | Very difficult | Somewhat difficult | Neither difficult nor easy | Somewhat easy | Very easy |
|---|---|---|---|---|---|
| The Tasso-SST device | ◯ | ◯ | ◯ | ◯ | ◯ |
| The blood stabilization tube | ◯ | ◯ | ◯ | ◯ | ◯ |

Did you experience any issues with the Tasso-SST blood collection device?

◯ Yes
◯ No

Please describe any issues you had with the Tasso-SST blood collection device. (Optional)

__________________________________________

Did you experience any issues with the stabilizer tube or mixing your samples?

◯ Yes
◯ No

Please describe any issues you had with the stabilizer tube or mixing your samples. (Optional)

__________________________________________

Please use the space for any other comments on your experience today collecting blood samples using the homeRNA blood kit. (Optional)

__________________________________________

### Smoke Exposure Sample Collection Survey

Please complete the survey below.

Thank you!

Please enter the sample code of the kit you have just finished using: __________________________________

The code will be located on the outside of the individual kit box and will have the format of XXXX - X?

Please indicate the date and time you collected your sample.

Date: __________________________________

Time: __________________________________

You should have been provided a temperature and humidity monitor in the first package we sent you. Using this monitor, please report the temperature and humidity below.

Temperature (Farenheit): __________________________________

Humidity (%): __________________________________

What is the current PM 2.5 reading on the PurpleAir indoor air monitor? __________________________________

What is the current PM 2.5 reading on the closest outdoor PurpleAir monitor? __________________________________

Have your medications changed since you last filled out the enrollment survey?
- ◯ Yes
- ◯ No

Please select from the following:
- ◯ I started taking a new medication
- ◯ I stopped taking a medication
- ◯ My medications have not changed since I last filled out a survey

Please describe the new medication: __________________________________________

Please describe which medication you stopped taking: __________________________________________

Has your address changed since you last filled out a survey?
- ◯ Yes
- ◯ No

If yes, what is the zipcode of your new address? __________________________________

Do you anticipate your address will change in the next 2 months?
◯ Yes
◯ No

**We will now ask you a few questions about your exposure to wildfire smoke in the past week:**

Approximately how many hours were you outside when it was smoky:

In the past week? ______________________

In the last 3 days? ______________________

In the last 24 hours? ______________________

Approximately how many hours did you spend indoors at home:

In the past week? ______________________

In the last 3 days? ______________________

In the last 24 hours? ______________________

Approximately how many hours did you spend indoors at work (if you do not work from home):

In the past week? ______________________

In the last three days? ______________________

In the last 24 hours? ______________________

Approximately how many hours did you spend in a car or bus:

In the past week? ______________________

In the last 3 days? ______________________

In the last 24 hours? ______________________

When you were outside, were you wearing a mask or any other sort of respiratory protection?
◯ Yes
◯ No

| | |
|---|---|
| What kind of protection were you using? | ◯ N95 respirator<br>◯ Surgical mask<br>◯ Cloth mask<br>◯ Other |
| If other, please specify: | __________________________________ |
| Did you use any sort of air filters or other mitigation strategies at home to improve indoor air quality? | ◯ Yes<br>◯ No |
| Select all mitigation strategies that you used this week. | ☐ I kept windows and doors closed<br>☐ I did not vacuum or dust<br>☐ I avoided cooking using a gas stove<br>☐ I used a box fan with an attached MERV filter<br>☐ I used a portable HEPA air filter<br>☐ I used the filter unit on my built in A/C ventilation system<br>☐ I avoided outdoor recreation or work<br>☐ I did something else |
| Please describe other mitigation strategies that you used this week. | __________________________________ |

**We will now ask you a few questions about how you are currently feeling:**

| | |
|---|---|
| Do you currently have any of these physical symptoms? | ☐ Coughing<br>☐ Trouble breathing normally<br>☐ Stinging eyes<br>☐ A scratchy throat<br>☐ Runny nose<br>☐ Irritated sinuses<br>☐ Wheezing and shortness of breath<br>☐ Chest pain<br>☐ Headaches<br>☐ Asthma attack<br>☐ Tiredness<br>☐ Fast heartbeat<br>☐ Other<br>☐ I do not have any of these physical symptoms |
| If other, please describe your symptoms: | __________________________________ |
| Are you currently feeling sick (e.g., cold or flu)? | ◯ Yes<br>◯ No |
| Please briefly describe your symptoms related to feeling sick. | __________________________________ |

| | I was experiencing less stress than normal | | I was experiencing about the same amount of stress as normal | | I was experiencing more stress than normal |
|---|---|---|---|---|---|
| Compared to your normal level of stress, how stressed were you today? | ◯ | ◯ | ◯ | ◯ | ◯ |

| | I generally have the same amount of stress as normal | | I am somewhat more stressed compared to normal | | I can become extremely stressed compared to normal |
|---|---|---|---|---|---|
| Compared to your normal level of stress, how would you say you react to decreased air quality in your physical environment? | ◯ | ◯ | ◯ | ◯ | ◯ |

| | I would feel very comfortable trying to use this kit during a natural disaster | | I would feel neutral about trying to use this kit during a natural disaster | | I would feel very uncomfortable trying to use this kit during a natural disaster |
|---|---|---|---|---|---|
| How comfortable do you think you would be using this kit in the midst of a natural disaster, assuming you are not in any immediate danger? (e.g. wildfire, earthquake, hazardous weather) | ◯ | ◯ | ◯ | ◯ | ◯ |

Apart from wildfire smoke exposure, in the past week were you exposed to any environmental stressors such as poor air quality, extreme temperatures, physical injuries, exposures to chemicals, etc.?

◯ Yes
◯ No

If yes, please specify:

__________________________________

Approximately how many hours of sleep did you get on average in the past week?

◯ Less than 6 hours
◯ 6 - 9 hours
◯ 9 - 12 hours
◯ More than 12 hours

Were you able to engage in your normal levels of exercise in the past week?

◯ Yes
◯ No
◯ I do not normally exercise

If not, please indicate any reasons you were unable to exercise, if applicable (optional):

__________________________________

| | |
|---|---|
| Were you able to engage in your normal levels of mindfulness activities in the past week? | ◯ Yes<br>◯ No<br>◯ I do not normally practice mindfulness |
| If not, please indicate any reasons you were unable to practice mindfulness, if applicable (optional): | __________________________________ |
| Have you been able to engage in your other hobbies this past week? (e.g., gardening, arts/crafts, video games, reading, writing, watching movies or TV, etc.) | ◯ Yes<br>◯ No<br>◯ I do not normally engage in other hobbies |
| If not, please indicate any reasons you were unable to engage in your other hobbies, if applicable (optional): | __________________________________ |

**We will now ask a few questions about your experience when using the homeRNA blood kit today:**

| | |
|---|---|
| Approximately how long (in minutes) did it take to use the homeRNA blood kit today? | ◯ Less than 5 minutes<br>◯ 5-10 minutes<br>◯ 11-15 minutes<br>◯ 16-20 minutes<br>◯ More than 20 minutes |
| While using the homeRNA blood kit today, approximately how long (in minutes) did you leave the Tasso-SST blood collection device on your arm? | ◯ Less than 3 minutes<br>◯ 3 - 5 minutes<br>◯ More than 5 minutes<br>◯ I am unsure |
| Based on the image below, approximately how much blood did you collect today? Choose the closest level. | ◯ I was unable to collect any blood<br>◯ Level 1<br>◯ Level 2<br>◯ Level 3<br>◯ Level 4 |

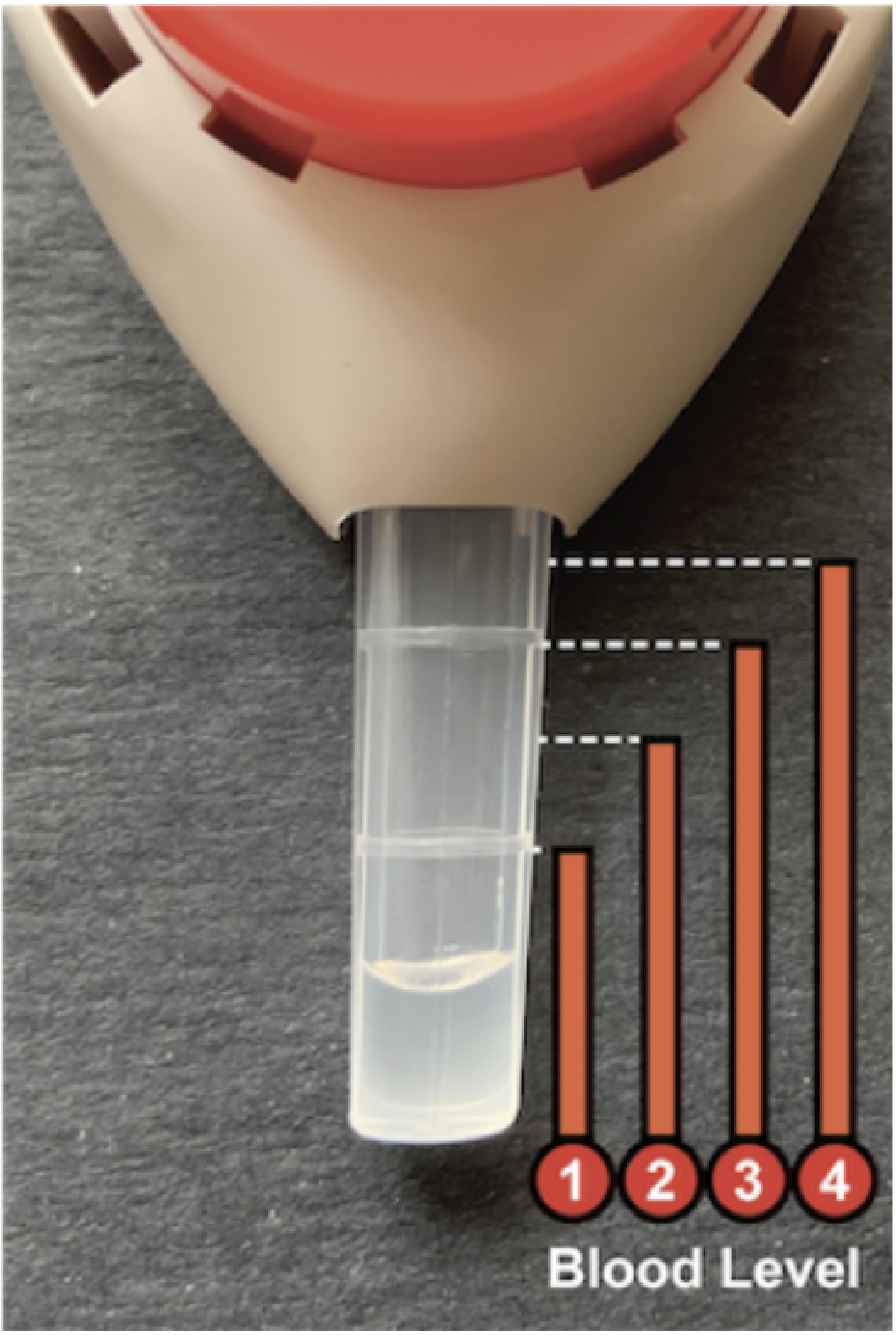


| Did you experience any pain while using the homeRNA blood kit today? | ◯ No pain<br>◯ Mild pain<br>◯ Moderate pain<br>◯ Severe pain<br>◯ Very severe pain |
|---|---|
| Has the temporary pain resolved? | ◯ Yes<br>◯ No |
| How long did the pain last? | ______________________________ |

**How easy was it to use the components of the kit listed below?**

| | Very difficult | Somewhat difficult | Neither difficult nor easy | Somewhat easy | Very easy |
|---|---|---|---|---|---|
| The Tasso-SST device | ○ | ○ | ○ | ○ | ○ |
| The blood stabilization tube | ○ | ○ | ○ | ○ | ○ |

Did you experience any issues with the Tasso-SST blood collection device?

○ Yes
○ No

Did you experience any issues with the stabilizer tube or mixing your samples?

○ Yes
○ No

Please use the space for any other comments on your experience today collecting blood samples using the homeRNA blood kit. (Optional)

__________________________________________

### Closing Survey

Please complete the survey below.

A $20 gift card link will be sent to your email after completing the survey.

Thank you!

**We will first ask some questions about events that may have changed either your smoke exposure or your immune response to smoke exposure over the last year.**

Did you travel anywhere in the past year where you were exposed to more or less wildfire smoke than if you had stayed home?

- ◯ Yes
- ◯ No

Please indicate the dates and city where you traveled.

__________________________________
(i.e., Chicago, IL from July 18-24, 2021)

Do you smoke or have you ever smoked?

- ◯ Yes, I currently smoke
- ◯ I do not currently smoke, but I have smoked before
- ◯ I have never smoked

In the past year, did any of the following significant life events occur? Select all that apply.

- ☐ Started or stopped smoking
- ☐ Became pregnant
- ☐ Received a new medical diagnosis
- ☐ Significant changes in diet or exercise
- ☐ Significant changes in sleep schedule
- ☐ Other life event that you would like to share with the study team (optional)
- ☐ No significant life events to report

Please briefly describe this new medical diagnosis (optional)

__________________________________

Please briefly describe the significant changes in diet or exercise (optional)

__________________________________

Please briefly describe the significant changes in sleep schedule (optional)

__________________________________

Please describe the other significant life event (optional)

__________________________________

Please indicate approximately what date(s) significant life event(s) occurred

__________________________________
(e.g., Stopped smoking mid-august 2021 and went on a Vegan Diet between February-March 2022)

#### We will now ask you a few questions about race and ethnicity.

Are you of Hispanic, Latino, or Spanish origin? Select all that apply.

- ☐ No, not Hispanic, Spanish, Latino, or Spanish origin
- ☐ Yes, Mexican, Mexican-American, Chicano
- ☐ Yes, Puerto Rican
- ☐ Yes, other Hispanic, Latino, or Spanish origin
- ☐ Prefer not to answer

If you selected other Hispanic, Latino, or Spanish origin please specify here (for example, Salvadoran, Dominican, Colombian, Guatemalan, Spaniard, Ecuadorian, etc.) (optional)

__________________________________

What is your race or origin? Select all that apply.

- ☐ American Indian or Alaskan Native
- ☐ Asian
- ☐ Black or African-American
- ☐ Native Hawaiian or Other Pacific Islander
- ☐ White, Middle Eastern or North African
- ☐ Other
- ☐ Prefer not to answer

If you selected American Indian or Alaskan Native above, please specify. Select all that apply. (optional)

- ☐ American Indian
- ☐ Alaskan Native
- ☐ Central or South American Indian
- ☐ Other American Indian or Alaskan Native
- ☐ Unknown

If you selected Other American Indian or Alaskan Native, please specify here. (optional)

__________________________________

If applicable, write the name(s) of enrolled or principal tribe(s), for example, Navajo Nation, Blackfeet Tribe, Mayan, Aztec, Native Village of Barrow Inupiat Traditional Government, Nome Eskimo Community, etc. (optional)

__________________________________

If you selected Asian above, please specify. Select all that apply. (optional)

- ☐ Chinese
- ☐ Filipino
- ☐ Asian Indian
- ☐ Vietnamese
- ☐ Japanese
- ☐ Other Asian
- ☐ Unknown

If you selected Other Asian above, please specify (for example, Pakistani, Cambodian, Hmong, Laotian, etc). (optional)

__________________________________

| | |
|---|---|
| If you selected Black or African-American above, please specify. Select all that apply. (optional) | ☐ African-American<br>☐ Caribbean<br>☐ Ethiopian<br>☐ Ghanaian<br>☐ Haitian<br>☐ Jamaican<br>☐ Liberian<br>☐ Nigerian<br>☐ Somali<br>☐ South African<br>☐ Other Black or African American<br>☐ Unknown |
| If you selected Other Black or African-American above, please specify. (optional) | ______________________ |
| If you selected Native Hawaiian or Other Pacific Islander above, please specify. Select all that apply. (optional) | ☐ Native Hawaiian<br>☐ Samoan<br>☐ Chamorro<br>☐ Other Pacific Islander<br>☐ Unknown |
| If you selected Other Pacific Islander above, please specify (for example, Tongan, Fijian, Marshallese, etc.). (optional) | ______________________ |
| If you selected White, Middle Eastern, or North African above, please specify. Select all that apply. (optional) | ☐ European<br>☐ Middle Eastern or North African<br>☐ Other White<br>☐ Unknown |
| If you selected European above, please specify if known. (for example, German, Irish, English, Italian, Polish, Scottish, Spanish, etc.). optional | ______________________ |
| If you selected Middle Eastern or North African above, please specify if known. (for example, Afghan, Algerian, Egyptian, Iranian, Iraqi, Israeli, Lebanese, Moroccan, Syrian, etc.). (optional) | ______________________ |
| If you selected Other White above, please specify if known. (optional) | ______________________ |
| If you selected other race or origin above, please specify if known (optional) | ______________________ |

**We will conclude with an opportunity for you to opt into receiving invitations to future research and community events from the study team.**

| | |
|---|---|
| We will be holding a community event in partnership with Clean Air Methow in the coming year. Would you like to receive an email invitation to this event?<br><br>Please note, if you select yes this does not mean you are committed to attending. Selecting yes means you will be sent an email with an invitation from the study team between?June and September of this year. | ◯ Yes<br>◯ No |

Would you like to be contacted if we are looking for participants to participate in a second year of this study for the 2022 wildfire season?
Please note, if you select yes this does not mean you are committed to participation in this study. Selecting yes will mean you will be sent an email with an invitation to sign up between May and July of this year. To participate in a second year of this study, you will need to sign a new consent form and fill out a new enrollment survey.

◯ Yes
◯ No

Do you have any feedback for the study team? (optional)

______________________________

### Follow Up Survey

| | |
|---|---|
| We sent one extra blood collection device with each shipment of three kits. How often did you use your extra Tasso-STT blood collection device?? | ◯ Always<br>◯ Often<br>◯ Sometimes<br>◯ Rarely<br>◯ Never<br>◯ I don't remember |
| Which of the following motivated you to enroll in the study (please check all that apply)? | ☐ An interest in helping my community understand the health impacts of wildfire smoke<br>☐ An interest in adding to the body of knowledge on the health impacts of wildfire smoke<br>☐ A general interest in science<br>☐ An interest in learning about the Tasso-STT blood collection device<br>☐ A friend or family member who was also participating in the study<br>☐ The financial compensation I received for participating in the study<br>☐ Other motivation not listed above |
| Please explain | __________________________________ |
| Have you participated in an in-person blood sampling study (e.g., having your blood drawn at a blood draw facility or clinic for the purpose of a research study) previously? | ◯ Yes<br>◯ No<br>◯ Unsure |
| We would like to understand how participating in a remote blood sampling study with multiple samples collected (the current study) compares to participating in an in-person blood sampling study (e.g., having your blood drawn at a blood draw facility or clinic). If you haven't participated in an in-person study previously, please imagine yourself in that scenario. Please rate the ease of participating in a study with remote blood sampling compared to participating in a study with in-person blood sampling.? | ◯ Remote blood sampling is significantly easier<br>◯ Remote blood sampling is somewhat easier<br>◯ Remote blood sampling is neither easier nor harder than in-person<br>◯ Remote blood sampling is somewhat harder<br>◯ Remote blood sampling is significantly harder |
| Would any of the following prevent you from participating in an in-person blood sampling study (please check all that apply)? | ☐ Length of commute to the nearest clinic or blood draw facility<br>☐ Difficulty obtaining transportation to the clinic or blood draw facility<br>☐ Difficulty fitting blood draw visits into a work schedule or caregiver schedule<br>☐ Discomfort with the method of drawing blood at clinics or blood draw facilities (e.g., using a syringe or collection needle to draw blood from the inner elbow)<br>☐ Discomfort with going to clinics or other medical facilities<br>☐ Other |
| Please explain | __________________________________ |

Would any of the following prevent you from participating in another remote blood sampling study (Please check all that apply)?

- ☐ Difficulty understanding the instructions for using the Tasso-STT blood collection device
- ☐ Difficulty taking a sample of your own blood
- ☐ Discomfort with the method of drawing blood at home if using a different method than used in this study (e.g., using a finger stick to draw blood)
- ☐ Difficulty taking multiple blood samples over the course of a week
- ☐ Difficulty participating in the study for the full duration (e.g., taking many blood samples over 6-9 months as you did in this study)
- ☐ Lack of privacy when taking a sample of your own blood if you didn't have a private space at home
- ☐ Other

Please explain

__________________________________

**Please rate your level of agreement with the following statements:**

| | 1 Strongly Disagree | 2 | 3 | 4 | 5 Strongly Agree |
|---|---|---|---|---|---|
| The instructions on how to use the Tasso-STT blood collection device within the kit were easy to follow. | ◯ | ◯ | ◯ | ◯ | ◯ |
| The instructions on how to use the stabilizer tube (the tube that you screw together with the blood tube) within the kit were easy to follow. | ◯ | ◯ | ◯ | ◯ | ◯ |
| The instructions and communications from the study team were easy to follow | ◯ | ◯ | ◯ | ◯ | ◯ |
| A phone call after I enrolled in the study that explained the steps of study participation would have been helpful. | ◯ | ◯ | ◯ | ◯ | ◯ |
| The flexibility in when I could take my blood sample (e.g., at the time of day that was most convenient) was an important factor in my ability to participate in this study. | ◯ | ◯ | ◯ | ◯ | ◯ |

In the current study, communication from the study team was sent to you over email. If you were to participate in the study again, how would you prefer to receive information for the study team?

- ◯ Email only (i.e., the same way you received information in this study)
- ◯ Text message only
- ◯ Email and text message
- ◯ Other

Please explain

__________________________________

| | |
|---|---|
| Did you watch the video on how to use the blood collection device and stabilization kit? | ◯ Yes<br>◯ No<br>◯ Unsure |
| The video was helpful. (1= strongly disagree, 5= strongly agree). | ◯ 1 Strongly agree<br>◯ 2<br>◯ 3<br>◯ 4<br>◯ 5 Strongly disagree |
| How comfortable do you think you would be using this kit in the midst of a natural disaster, assuming you are not in any immediate danger? (e.g. wildfire, earthquake, hazardous weather) (1= very uncomfortable, 5= very comfortable). | ◯ 1 Very uncomfortable<br>◯ 2<br>◯ 3<br>◯ 4<br>◯ 5 Very comfortable |
| Would you be willing to participate in this study or a similar study again? | ◯ Yes<br>◯ No<br>◯ Unsure |
| How could we improve this study in the future? | __________________________________ |
| Please indicate the maximum number of years that you would be willing to participate in this study or a similar remote blood sampling study: | ◯ 1 years<br>◯ 2 years<br>◯ 3 years<br>◯ 4 years<br>◯ 5 years |
| Is there anything we could change so that you would want to participate in this study or a similar remote blood sampling study again? | __________________________________ |
| Are there any other thoughts that you would like to share with the study team? | __________________________________ |
